# Cerebral Spinal Fluid Volumetrics and Paralimbic Predictors of Executive Dysfunction in Congenital Heart Disease: A Machine Learning Approach Informing Mechanistic Insights

**DOI:** 10.1101/2023.10.16.23297055

**Authors:** Vince K. Lee, Julia Wallace, Benjamin Meyers, Adriana Racki, Anushka Shah, Nancy H. Beluk, Laura Cabral, Sue Beers, Daryaneh Badaly, Cecilia Lo, Ashok Panigrahy, Rafael Ceschin

**Author notes:** The authors declare that the research was conducted in the absence of any commercial or financial relationships that could be construed as a potential conflict of interest. Data Sharing: Individual participant data that underlie the results reported in this article, after deidentification (text, tables, figures, and appendices), Study Protocol, and Statistical Analysis Plan, are available upon formal request. Requests should be submitted to the corresponding author. Corresponding Author: Rafael Ceschin, Ph.D. Department of Radiology, Children’s Hospital of Pittsburgh of UPMC 4401 Penn Ave., Pittsburgh, PA 15224.

## Abstract

The relationship between increased cerebral spinal fluid (CSF) ventricular compartments, structural and microstructural dysmaturation, and executive function in patients with congenital heart disease (CHD) is unknown. Here, we leverage a novel machine-learning data-driven technique to delineate interrelationships between CSF ventricular volume, structural and microstructural alterations, clinical risk factors, and sub-domains of executive dysfunction in adolescent CHD patients. We trained random forest regression models to predict measures of executive function (EF) from the NIH Toolbox, the Delis-Kaplan Executive Function System (D-KEFS), and the Behavior Rating Inventory of Executive Function (BRIEF) and across three subdomains of EF – mental flexibility, working memory, and inhibition. We estimated the best parameters for the random forest algorithm via a randomized grid search of parameters using 10-fold cross-validation on the training set only. The best parameters were then used to fit the model on the full training set and validated on the test set. Algorithm performance was measured using root-mean squared-error (RMSE). As predictors, we included patient clinical variables, perioperative clinical measures, microstructural white matter (diffusion tensor imaging- DTI), and structural volumes (volumetric magnetic resonance imaging- MRI). Structural white matter was measured using along-tract diffusivity measures of 13 inter-hemispheric and cortico-association fibers. Structural volumes were measured using FreeSurfer and manual segmentation of key structures. Variable importance was measured by the average Gini-impurity of each feature across all decision trees in which that feature is present in the model, and functional ontology mapping (FOM) was used to measure the degree of overlap in feature importance for each EF subdomain and across subdomains. We found that CSF structural properties (including increased lateral ventricular volume and reduced choroid plexus volumes) in conjunction with proximate cortical projection and paralimbic-related association white matter tracts that straddle the lateral ventricles and distal paralimbic-related subcortical structures (basal ganglia, hippocampus, cerebellum) are predictive of two-specific subdomains of executive dysfunction in CHD patients: cognitive flexibility and inhibition. These findings in conjunction with combined RF models that incorporated clinical risk factors, highlighted important clinical risk factors, including the presence of microbleeds, altered vessel volume, and delayed PDA closure, suggesting that CSF-interstitial fluid clearance, vascular pulsatility, and glymphatic microfluid dynamics may be pathways that are impaired in CHD, providing mechanistic information about the relationship between CSF and executive dysfunction.

## 1. Introduction

Cerebrospinal fluid (CSF) volume increase is a common finding in brains of fetuses^1,2^ and infants with congenital heart disease (CHD)^3–7^. Increased CSF volumes have been associated with poor cognitive, language, and motor skills^4,6^ as well as poor behavioral state regulation in neonates, infants, and young children with CHD^8^. Beyond the early childhood years, there is a paucity of research investigating the subsequent fate of these elevated CSF volumes and their potential associations with impairments in neurodevelopment during late childhood and adolescence within CHD. The few studies between brain structures and neurodevelopmental outcomes in adolescents with CHD excluded CSF^9^ or used a simplified single intraventricular volume for CSF ^10^.

Neurodevelopmental impairment is one of the major comorbidities in CHD, and in adolescence, one of its critical manifestations is poor executive function ^11,12^. Executive function (EF) encompasses a set of advanced cognitive skills essential for goal-oriented actions, self-regulation, and adaptability. Although there are different models of EF, Diamond’s seminal model identified three core domains: cognitive flexibility (adapting to challenges), inhibitory control (maintaining self-control), and working memory (quantity of information held and manipulated at a time).^13^ Deficits in these domains can adversely impact children’s development and adaptive functioning, with long-term consequences like lower academic achievement, increased reliance on remedial education, social adaptation difficulties, and reduced overall quality of life. Studies have explored factors and mechanisms potentially contributing to executive function impairment, including physiological aspects of heart lesion type, clinical care (surgical methods and medical management), and sociodemographic factors. Brain structures have also been studied: from broadly assessed brain hypoplasia and dysplasia^3^, to more refined volumes^14^ and cortical thickness and to more focal white matter microstructure^15^. However, the association between brain structural abnormalities and outcomes were highly variable, including instances of no significant associations^11,16^, and most of the comprehensive brain structural studies were in younger children or neonates.

The goal of this study was to examine CSF volume as neuroimaging features and their role in predicting EF impairments among adolescents with CHD while accounting for other brain structures as well as clinical and sociodemographic factors. However, due to the large number of brain structural, clinical, and sociodemographic factors involved, traditional methods for feature importance and covariate selection were not adequate. Furthermore, CSF and other brain structural volumes occupy the same well-defined cranial space, therefore the volumes may not be independent of each other. Therefore, we used explainable machine learning method, specifically, random forest regression. Random forest regression, in comparison to conventional methods, exhibits enhanced robustness in handling collinearity and outliers, and excels at revealing interactions among exposure variables. In this study, we used ensemble random forest regression to study the interplay between CSF volumes, brain macro- and microstructural measures, and sociodemographic/medical risk factors in relationship to EF in pediatric/adolescent CHD patients.

## 2. Methods

### 2.1. Participants

Participants were recruited from a single center, using print and digital advertisements, an online registry of healthy volunteers, and referrals within targeted cardiology outpatient clinics. Study exclusion criteria included comorbid genetic disorders, contraindications for MRI (e.g., a pacemaker), and non-English speakers. For healthy controls, study exclusion criteria also included preterm birth and neurological abnormalities (e.g., brain malformations, strokes, hydrocephalus). We initially screened 143 patients with CHD and 98 healthy controls (Supplemental Figure 1). A total of 69 CHD and 92 healthy controls underwent brain MR scanning. After removing cases with un-analyzable DTI due to motion artifact or technical factors, and only including participants with analyzable structural 3D T1 imaging, diffusion tensor imaging and EF measures, the final sample of patients included the following: CHD patients (n=57) and age-matched controls (n=86), with ages ranging from 6-17 years. These patients were prospectively recruited at our institution with Institutional Review Board (IRB) approval and oversight (for reference: University of Pittsburgh Institutional Review Board STUDY20060128: Multimodal Connectome Study approval 23 July 2020 and STUDY1904003 Ciliary Dysfunction, Brain Dysplasia, and Neurodevelopmental Outcome in Congenital approval 6 February 2023). The project was completed in accordance with the ethical principles of the Helsinki Declaration and parent/guardian consent was obtained. We have previously published portions of this prospectively recruited cohort.^17–21^ Patients with CHD included a heterogenous mix of cardiac lesions, including hypoplastic left heart syndrome (HLHS), aortic arch abnormalities, d-transposition of the great aorta (d-TGA), and other malformations requiring surgical correction in the first year of life. Clinical and surgical history from birth, medical risk factors, and additional social determinants of health (SDOH) were collected from the medical record (Table 1 and Supplemental Table 2).

### 2.2. Executive Function

As previously described^18^, executive function (EF) outcomes were measured using standardized cognitive tasks from the NIH Toolbox, the Delis-Kaplan Executive Function System (D-KEFS), the WISC-IV as well as parent-completed ratings from the Behavior Rating Inventory of Executive Function (BRIEF-2) (Table 1B). We considered four subdomains of EF: cognitive flexibility, working memory, inhibition, planning and problem-solving. We used age-corrected standard scores from the NIH Toolbox (*M* = 100; *SD* = 15), scaled scores from the D-KEFS (*M* = 10; *SD* = 3), and T-scores from the BRIEF (*M* = 50; *SD* = 10),

### 2.3. Image Acquisition

Participants underwent brain MRI on a 3 Tesla Skyra scanner (Siemens, Erlangen, Germany), using a 32-channel head coil. 3D sagittally acquired 42 directions images DTI and T1 images were used for our analysis. The DTI sequence had the following parameters: FOV = 256mm, voxel size = 2.0mm (isotropic), TE/TR=92ms/12600 ms, and 42-directions at B=1000s/mm2. The T1 sequence had the following parameters: TR = 2400 ms, TE = 3.16 ms, TI = 1200 ms, flip angle = 8°; 1 mm isotropic voxel size.

### 2.4. Image Processing and Feature Construction

Structural white matter features were generated using our previously developed automated tractography pipeline.^22^ Initially developed for group analysis of neonatal diffusion imaging, we have since generalized it to use with population and age-appropriate region of interest templates for tracking. We generated along-tract diffusivity measures – fractional anisotropy (FA), axial diffusivity (AD), radial diffusivity (RD), and mean diffusivity (MD) – of 18 inter-hemispheric and cortico-association fibers. The following tracts were used: left and right arcuate fasciculi, body, genu, and splenium of the corpus callosum, left and right corticospinal tracts, left and right cingulum, left and right fronto-occipital fasciculi, fornix, left and right inferior longitudinal fasciculi, left and right superior longitudinal fasciculi, and left and right uncinate fasciculi. As along-tract analysis generates several correlated measures for each tract (proportional to the length of the tract), we performed a data reduction step by aggregating along-tract values into quartiles along the primary direction of each tract (i.e., the first quartile of the corticospinal tract spans from the brainstem to level of the hypothalamus).

Structural grey matter features were generated using the standard FreeSurfer cortical and subcortical volume parcellations,^23^ with manual inspection of intermediate steps and control points added when necessary. Additional manual inspection and correction (when necessary) of the hippocampus and amygdala was performed for each participant (as these are key structures that are prone to segmentation errors due to mis-registration or motion artifacts), as well as further manual subdivision of the cerebellar vermis. Finally, as a measure of total tissue volumes, we used FSL’s FAST Segmentation to extract total CSF, total white matter, and total grey matter volumes from the structural T1 images.

### 2.5. Modeling Executive Function Outcomes with Random Forest Regression

As our primary goal was to identify the prognostic utility of imaging features with a targeted investigation of CSF-related features on predicting EF outcomes, we chose an implicitly interpretable machine learning algorithm – Random Forest Regression (RFR) – to model each outcome. Random Forest is an ensemble learning method that operates by constructing a large pre-determined number of decision trees. The more trees in the ‘forest’, the more robust the model becomes. Importantly, each decision tree in the model is trained on a subset of participants (sampled with replacement) and a subset of the available features. This generates robust models with built-in bootstrapped sampling that have been shown to generalize better than more ‘advanced’ algorithms.^24^ Though Random Forest models are generally regarded as black-box models due to their complex ensemble nature, they do possess an implicit interpretability via the Gini coefficient (or Gini impurity) – the measure used to optimally split each node in the decision tree. A feature that provides a split which most reduces Gini impurity in the resulting child nodes is considered important for that split. By aggregating this information across all trees in the random forest, we can estimate an overall metric of feature importance across the model by observing which features are often used to split nodes, and by how much they can decrease the impurity.

We measured the contribution of imaging-derived features to our models’ predictive accuracy by generating two baseline models: 1) RFR model using only neuroimaging-derived features 2) RFR model using only clinical and surgical risk factors combined with SDOH features. We then compared the performance of each baseline model to 3) a full combined model (all available features) as a measure of information gain of each feature set. Figure 1 shows an overview of the model building and hyperparameter selection for each outcome. A separate RFR model was optimized and trained for each outcome measure using the Python Scikit-Learn library.^25^ Each independent dataset was partitioned into 80% for training and 20% for testing. We estimated the best parameters for the random forest algorithm via a randomized grid search of parameters using 10-fold cross-validation on the training set only. The best parameters were then used to fit the model on the full training set and validated on the test set. Algorithm performance was measured using root-mean squared-error (RMSE). A model was considered to be performing within an acceptable error margin if the RMSE in the test set was below one standard deviation of the normed parameters for each outcome. Variable importance was measured by the average Gini-impurity of each feature across all decision trees in which that feature is present in the model.

**Figure 1A.**
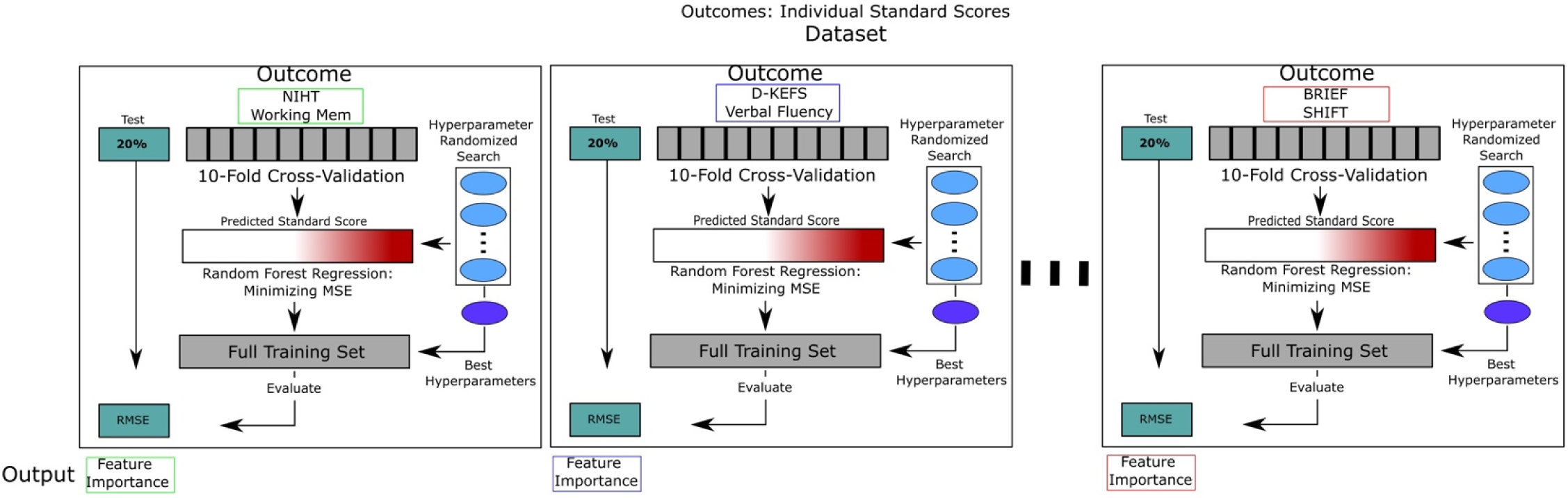
Hyperparameter selection strategy for each outcome. A separate RFR model was trained for each outcome measure for each dataset. Each independent dataset was partitioned into 80% for training and 20% for testing. We estimated the best parameters for the random forest algorithm via a randomized grid search of parameters using 10-fold cross-validation on the training set only. The best parameters were then used to fit the model on the full training set and validated on the test set. Algorithm performance was measured using root-mean squared-error (RMSE).

**Figure 1B.**
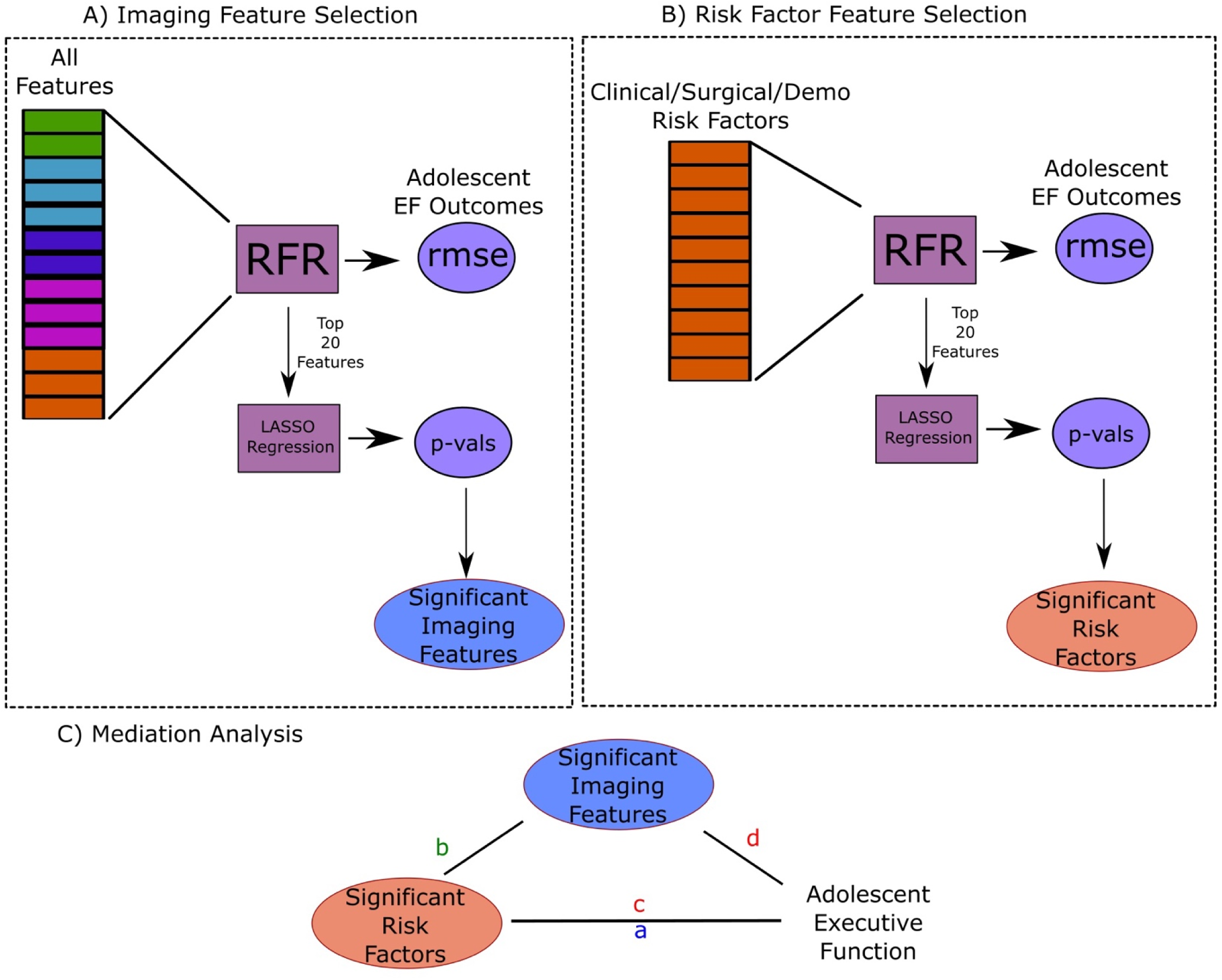
Feature selection schema for mediation analysis. Model selection filtered in models that A) passed the selection criteria for acceptable error margin (within 1 sd of normed test) and B) had important CSF features in the imaging features only models. From these models, we selected the main effects as A) the top 5 features in the SDOH/Clinical/Surgical models and B) any additional features selected via LASSO regression in the combined models. The mediator selection was A) CSF features in the top 20 imaging models and B) any additional imaging derived feature selected via LASSO regression in either the imaging only or combined models.

### 2.6. Statistical Validation and Mediation Analysis

A secondary goal of this work is a mechanistic exploration of the mediating effect of CSF imaging features identified as important for model performance on the direct effect of surgical, clinical, and SDOH features on EF outcomes. Due to the large number of features in the initial model, we implemented a data-driven feature selection approach. Although feature importance via Gini-coefficient provides an interpretable and robust measure for feature selection, it is not a sufficient criterion for selection alone due to its stochastic properties, and requires further statistical confirmation. We first used the ranked RFR features to select the top 20 important features from each model. We measured the total variance explained (R^2^) by the top features using ordinary least squares (OLS) regression. As many of the features used in the model are highly correlated, we then used Least Absolute Shrinkage and Selection Operator (LASSO) regularized regression with a regularization parameter (λ) of 0.1 to further select for a subset of features that are statistically significantly predictive of the outcome (when in the presence of correlated features). LASSO employs L1 regularization which adds the absolute values of the coefficients multiplied by the regularization parameter to the loss function. This additional constraint on the model parameters mitigates multicollinearity, and is an efficient method for variable selection. In the specific context of correlated features, given a group of correlated variables, LASSO tends to select one variable and shrink the remaining within-group coefficients towards zero, while minimizing loss in variance explained.

Our final selection criteria for potential direct effects and mediators are as follows. First, we only selected models that A) passed the selection criteria for acceptable error margin (within 1 sd of normed test) and B) had important CSF features in the imaging features only models. From these models, we selected the main effects as A) the top 5 features in the SDOH/Clinical/Surgical models and B) any additional features selected via LASSO regression in the combined models. The mediator selection was A) CSF features in the top 20 imaging models and B) any additional imaging derived feature selected via LASSO regression in either the imaging only or combined models.

Figure 2 shows the concept for the mediation analyses. The mediation analyses were performed by setting each executive function outcome (EF) as the dependent variable, the mediator as the identified imaging-derived feature, and the independent variables the identified clinical/surgical/SDOH risk factor. We constructed three sets of regression models: 1) the total effect of Risk Factor (RF) on EF: EF = b0 + a*RF + e; 2) the effect of RF on the mediator Imaging Feature (only a mediator if we detect a significant effect on b): IF = b0 +b*RF + e; 3) If b is significant, we then model the mediating effect of the IF over RF on EF: EF = b0 + c*RF + d*IF + e. If d (mediating effect) is significant, and c (direct effect) is a smaller effect than a, IF is a (partial) mediator of LRF on EF. All statistical analyses were performed using the Python statsmodels package.^26^

**Figure 2.**
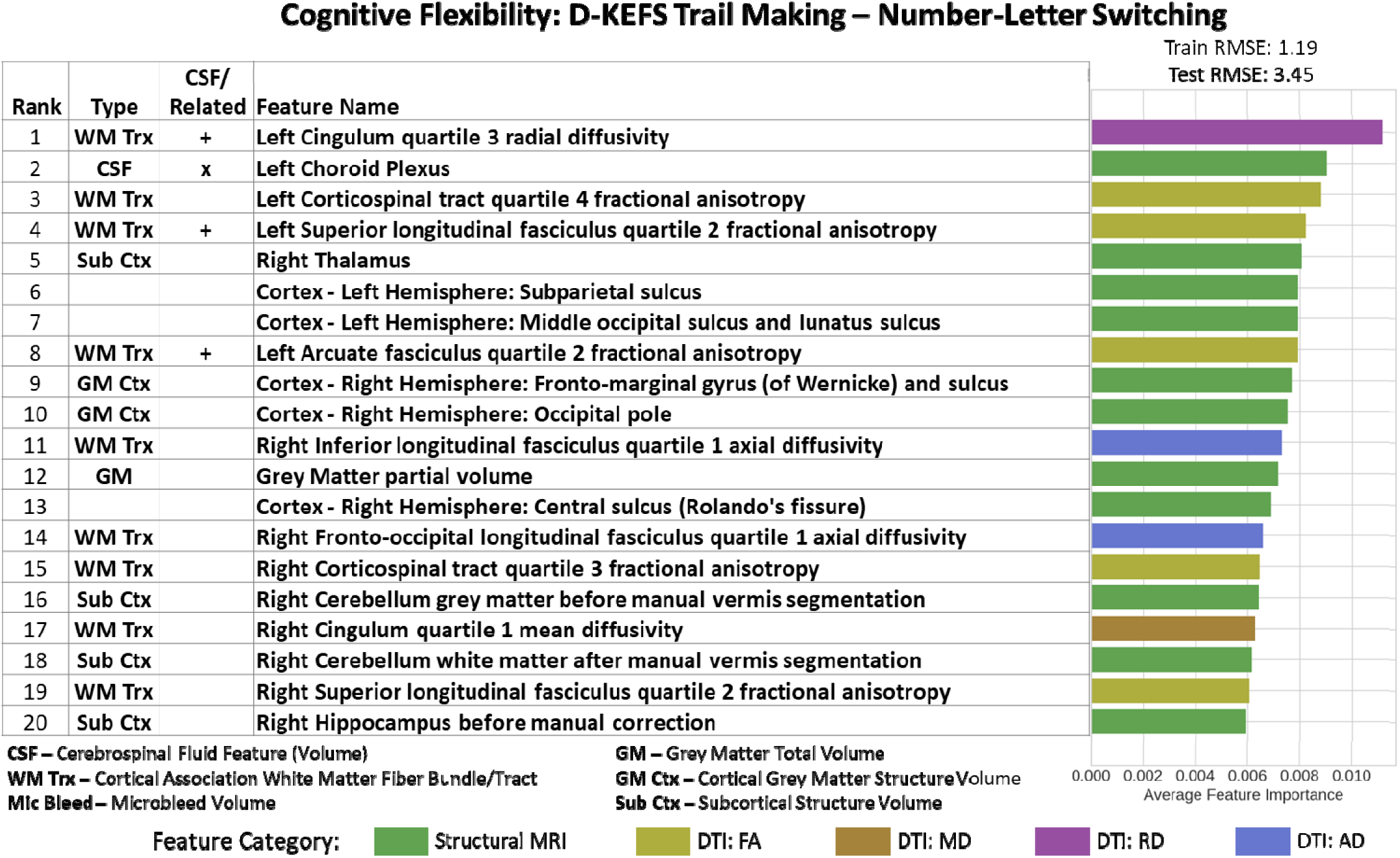
Imaging Only Top 20 Feature Importance results for Cognitive Flexibility Domain assessed using D-KEFS Trail Making – Number-Letter Switching Test. Left Choroid Plexus volume (rank 2) was the important CSF feature in this model along with proximate projection and association white matter tracts that straddle the left lateral ventricle, including the left cingulum (paralimbic cortical association-radial diffusivity), the left cortical spinal tract (primary motor projection-fractional anisotropy), the left superior longitudinal fasciculus-(cortical association-fractional anisotropy) and the left arcuate fasciculus (cortical association fibers-fractional anisotropy), selected cortical grey matter structures, and more distal contralateral subcortical structures (right thalamus, right cerebellum and right hippocampus).

### 2.7. Feature Ontology Mapping

There are two interpretability limitations to our methods thus far: 1) feature importance measures derived from random forest models are not deterministic, therefore a direct measure of feature importance rankings is not feasible; 2) highly correlated features – both quantitative and within similar functional groupings – may show artificially reduced significance when combined in a large model. To address these limitations, we performed Feature Ontology Mapping (FOM), which refers to systematically categorizing and selecting features based on their semantic relationships and shared characteristics to provide a deeper understanding of the relationships between the features and their effects on the target variable.^27,28^ Mapping features to an ontology allows the integration of domain expertise into the model, and also allows for validation of model decisions against domain knowledge, checking whether the model’s emphasis on certain features aligns with expert understanding.

We mapped each feature into an ontology of functional domains (i.e., motor, language, paralimbic, relay, etc.) and measured each functional domain’s contribution to feature importance for each outcome measure. See Supp. Table 4 for the full ontological mapping of features to functional domains. To account for disproportionate representation of different domains in the feature set, we measured the domain importance using two metrics: Domain Absolute Importance (% of functional domain represented in Top X Features) and Domain Relative Importance (% of functional domain represented in Top X Features / % of functional domain Represented in Total Features).

## RESULTS

There were no differences in gender incidence or age at MRI scan/EF measures between the CHD cohort and the healthy control cohort. While the level of maternal education was decreased in the CHD cohort compared to the healthy control cohort, there was no difference in the Child Opportunity Index between the two cohorts (Table 1A). Deficits in certain subdomains of executive function were noted in the CHD cohort compared to the healthy control cohort (Table 1B).

**Table 1A.**
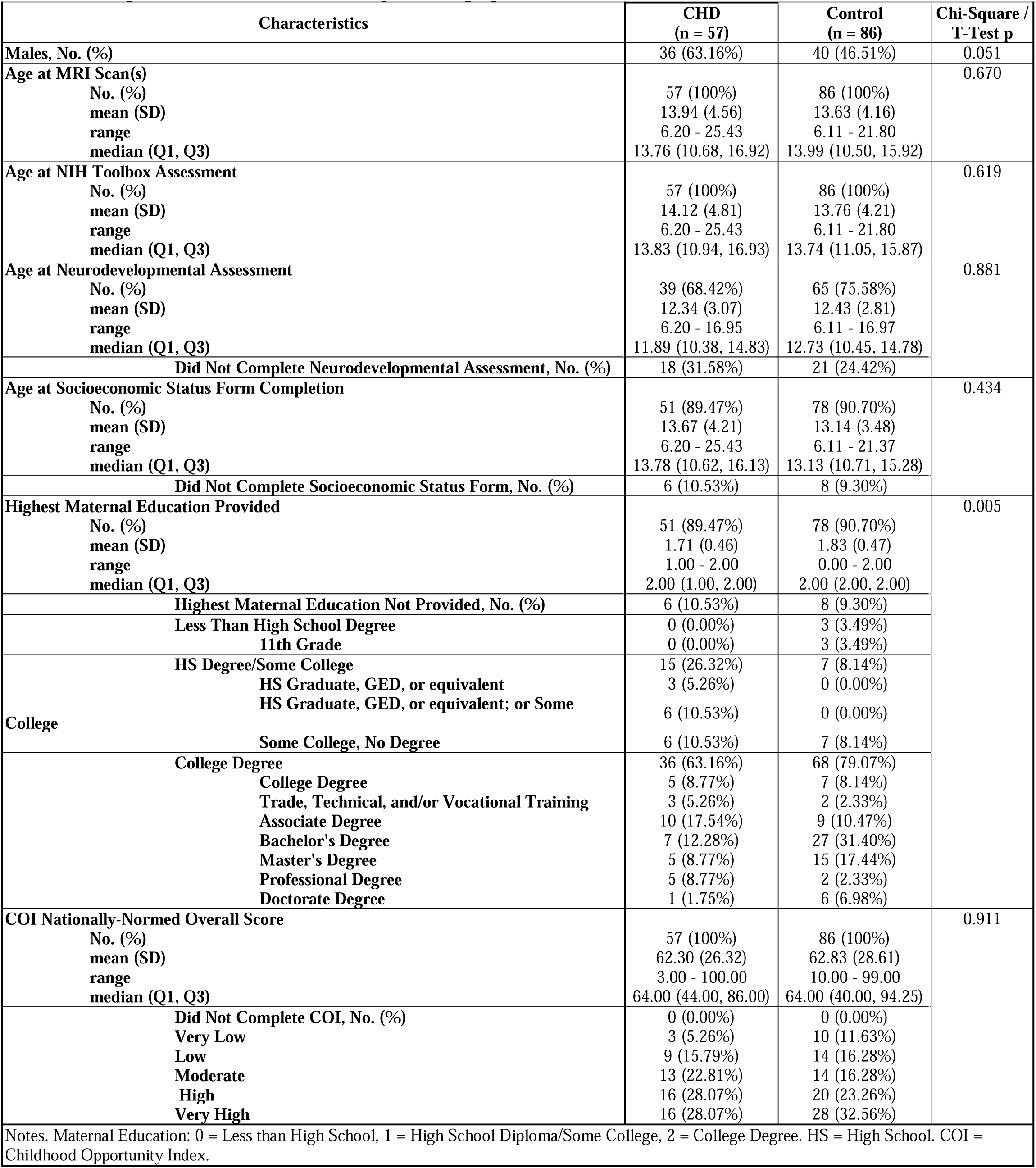
Comparison of CHD vs. Control Subject Demographics.

**Table 1B.** Comparison of CHD vs. Control Executive Function Neurocognitive Outcomes.

| Neurodevelopmental Assessment Outcomes | CHD<br>(n = 57) | Control<br>(n = 86) | T-Test p |
| --- | --- | --- | --- |
| **NIH Toolbox Cognitive Battery Age at Completion, years**<br>No. (%)<br>mean (SD)<br>range<br>median (Q1, Q3) | 57 (100%)<br>14.12 (4.81)<br>6.20 - 25.43<br>13.83 (10.94, 16.93) | 86 (100%)<br>13.76 (4.21)<br>6.11 - 21.80<br>13.74 (11.05, 15.87) | 0.619 |
| **Dimensional Card Change Sort Test, age corrected standard score**<br>No. (%)<br>mean (SD)<br>range<br>median (Q1, Q3) | 57 (100%)<br>95.57 (17.07)<br>66.49 - 148.00<br>96.00 (82.00, 105.00) | 86 (100%)<br>102.19 (15.14)<br>71.00 - 145.00<br>102.92 (91.74, 112.00) | 0.016 |
| **List Sorting Working Memory Test, age corrected standard score**<br>No. (%)<br>mean (SD)<br>range<br>median (Q1, Q3) | 56 (98.25%)<br>102.55 (14.34)<br>66.93 - 135.00<br>101.90 (92.89, 111.42) | 86 (100%)<br>106.17 (13.98)<br>80.00 - 147.00<br>106.11 (97.89, 114.75) | 0.137 |
| **Flanker Inhibitory Control & Attention Test, age corrected standard score**<br>No. (%)<br>mean (SD)<br>range<br>median (Q1, Q3) | 57 (100%)<br>96.23 (14.92)<br>66.00 - 128.00<br>96.00 (84.00, 105.09) | 86 (100%)<br>100.49 (13.75)<br>67.00 - 138.00<br>102.13 (89.83, 109.76) | 0.082 |
| **NIH Toolbox Sensation Battery Age at Completion, years**<br>No. (%)<br>mean (SD)<br>range<br>median (Q1, Q3) | 56 (98.25%)<br>14.08 (4.51)<br>6.20 - 25.43<br>13.82 (10.87, 16.93) | 86 (100%)<br>13.76 (3.97)<br>6.11 - 21.80<br>13.74 (11.05, 15.87) | 0.658 |
| **Odor Identification, age corrected standard score**<br>No. (%)<br>mean (SD)<br>range<br>median (Q1, Q3) | 56 (98.25%)<br>95.60 (13.46)<br>58.00 - 121.00<br>94.50 (89.09, 104.84) | 85 (98.84%)<br>95.39 (16.32)<br>64.00 - 132.15<br>93.00 (84.00, 107.00) | 0.937 |
| **D-KEFS Age at Completion, years**<br>No. (%)<br>mean (SD)<br>range<br>median (Q1, Q3) | 35 (61.40%)<br>12.83 (2.74)<br>8.09 - 16.95<br>13.05 (10.99, 15.18) | 58 (67.44%)<br>13.06 (2.23)<br>8.25 - 16.97<br>12.98 (11.51, 15.01) | 0.657 |
| **Trail Making - Condition 4: Number-Letter Switching, scaled score**<br>No. (%)<br>mean (SD)<br>range<br>median (Q1, Q3) | 32 (56.14%)<br>8.53 (2.72)<br>2.00 - 15.00<br>9.00 (7.00, 10.00) | 57 (66.28%)<br>11.00 (2.24)<br>4.00 - 15.00<br>11.00 (10.00, 12.00) | <0.001 |
| **Verbal Fluency - Condition 3: Category Switching - Correct Responses, scaled score**<br>No. (%)<br>mean (SD)<br>range<br>median (Q1, Q3) | 34 (59.65%)<br>10.03 (2.53)<br>5.00 - 17.00<br>10.00 (8.25, 11.75) | 58 (67.44%)<br>11.02 (3.31)<br>3.00 - 19.00<br>11.00 (9.00, 14.00) | 0.111 |
| **Verbal Fluency - Condition 3: Category Switching - Switching Accuracy, scaled score**<br>No. (%)<br>mean (SD)<br>range<br>median (Q1, Q3) | 34 (59.65%)<br>10.62 (2.39)<br>6.00 - 17.00<br>11.00 (9.00, 12.00) | 58 (67.44%)<br>11.02 (3.25)<br>3.00 - 19.00<br>11.00 (9.00, 13.00) | 0.501 |
| **Color Word Interference - Condition 3: Inhibition, scaled score**<br>No. (%)<br>mean (SD)<br>range<br>median (Q1, Q3) | 25 (43.86%)<br>9.04 (2.99)<br>3.00 - 14.00<br>10.00 (8.00, 11.00) | 55 (63.95%)<br>10.58 (2.71)<br>1.00 - 15.00<br>11.00 (9.00, 12.00) | 0.025 |
| **WISC-IV\* Age at Completion, years** |  |  | 0.940 |
| No. (%) | 39 (68.42%) | 64 (74.42%) |  |
| mean (SD) | 12.34 (3.07) | 12.39 (2.81) |  |
| range | 6.20 - 16.95 | 6.11 - 16.97 |  |
| median (Q1, Q3) | 11.89 (10.38, 14.83) | 12.70 (10.39, 14.67) |  |
| **Digit Span, scaled score** |  |  | 0.026 |
| No. (%) | 39 (68.42%) | 64 (74.42%) |  |
| mean (SD) | 9.23 (2.77) | 10.56 (2.96) |  |
| range | 4.00 - 16.00 | 4.00 - 19.00 |  |
| median (Q1, Q3) | 10.00 (7.00, 11.00) | 11.00 (8.00, 12.00) |  |
| **BRIEF-2 Parent Report Age at Completion, years** |  |  | 0.881 |
| No. (%) | 39 (68.42%) | 65 (75.58%) |  |
| mean (SD) | 12.34 (3.07) | 12.43 (2.81) |  |
| range | 6.20 - 16.95 | 6.11 - 16.97 |  |
| median (Q1, Q3) | 11.89 (10.38, 14.83) | 12.73 (10.45, 14.78) |  |
| **Shift, T-score** |  |  | 0.013 |
| No. (%) | 39 (68.42%) | 65 (75.58%) |  |
| mean (SD) | 50.54 (12.79) | 44.72 (7.71) |  |
| range | 36.00 - 88.00 | 36.00 - 66.00 |  |
| median (Q1, Q3) | 47.00 (40.00, 57.50) | 41.00 (38.00, 50.00) |  |
| **Working Memory, T-score** |  |  | 0.002 |
| No. (%) | 39 (68.42%) | 65 (75.58%) |  |
| mean (SD) | 56.36 (13.83) | 48.42 (9.67) |  |
| range | 35.00 - 82.00 | 36.00 - 78.00 |  |
| median (Q1, Q3) | 56.00 (43.00, 67.00) | 45.00 (40.00, 53.00) |  |
| **Inhibit, T-score** |  |  | 0.028 |
| No. (%) | 39 (68.42%) | 65 (75.58%) |  |
| mean (SD) | 49.72 (9.50) | 45.89 (6.04) |  |
| range | 36.00 - 86.00 | 37.00 - 66.00 |  |
| median (Q1, Q3) | 48.00 (44.50, 53.00) | 44.00 (42.00, 48.00) |  |
| **Planning & Organization, T-score** |  |  | <0.001 |
| No. (%) | 39 (68.42%) | 65 (75.58%) |  |
| mean (SD) | 54.82 (12.33) | 46.92 (9.80) |  |
| range | 33.00 - 84.00 | 33.00 - 78.00 |  |
| median (Q1, Q3) | 53.00 (46.50, 62.00) | 43.00 (40.00, 50.00) |  |
| * = 2 subjects assessed with WISC-V. Abbreviations: D-KEFS = Delis-Kaplan Executive Function System. WISC-IV = Wechsler Intelligence Scale for Children Fourth Edition. BRIEF-2 = Behavior Rating Inventory of Executive Function Second Edition. |  |  |  |

### 3.1 Random Forest Neuroimaging Model of Executive Function

Every imaging feature – from macrostructure regional volumes and global whole brain tissues volumes to microstructure white matter bundle assessments – was included in the imaging only random forest regression model for feature importance testing. The full list of neuroimaging features used in the models is listed in Supplemental Table 1: Neuroimaging Features (n= 495). The summary of feature importance results from the neuroimaging random forest regression models are presented in Table 2.

**Table 2.**
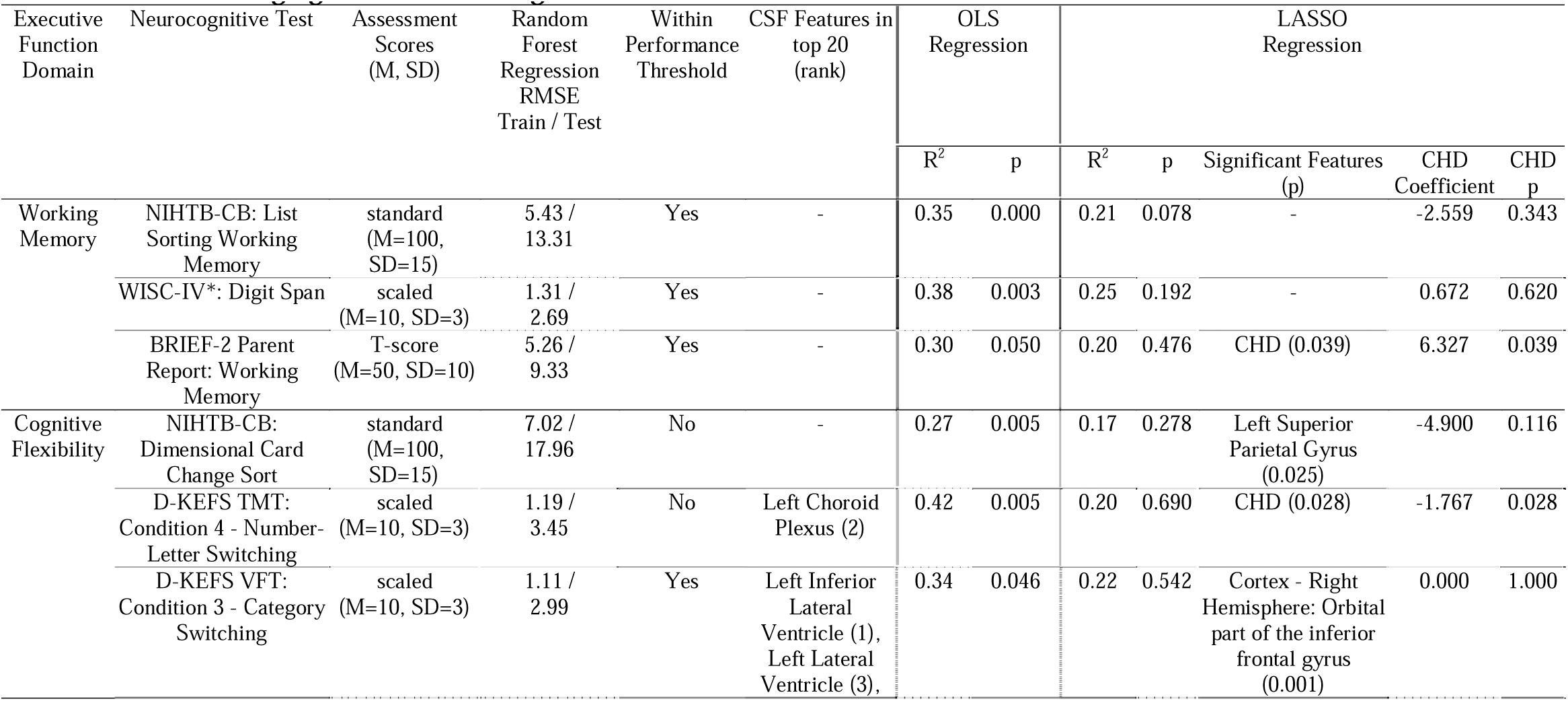
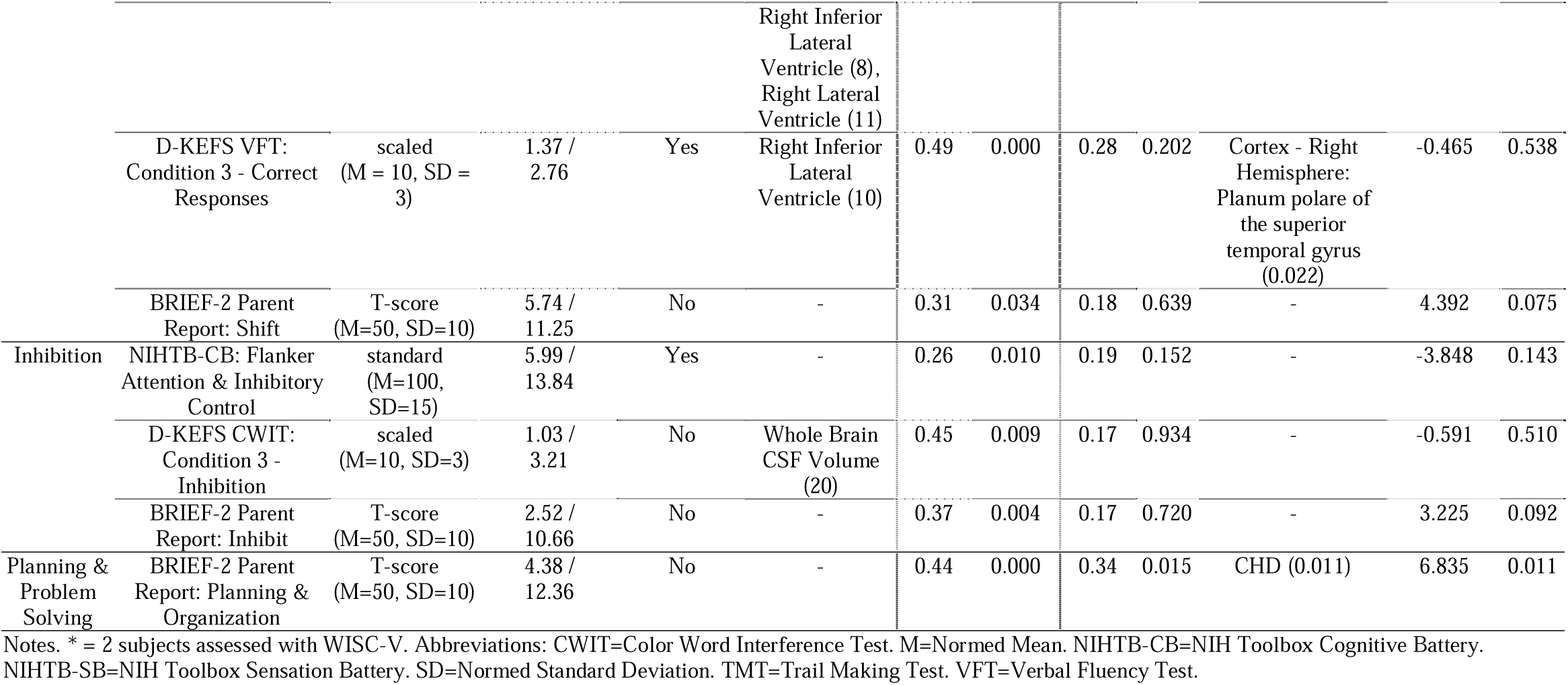
RF Neuroimaging Model Based Regressions.

| Executive Function Domain | Neurocognitive Test | Assessment Scores (M, SD) | Random Forest Regression RMSE Train / Test | Within Performance Threshold | CSF Features in top 20 (rank) | OLS Regression |  | LASSO Regression |  |  |  |  |
| --- | --- | --- | --- | --- | --- | --- | --- | --- | --- | --- | --- | --- |
|  |  |  |  |  |  | R<sup>2</sup> | p | R<sup>2</sup> | p | Significant Features (p) | CHD Coefficient | CHD p |
| Working Memory | NIHTB-CB: List Sorting Working Memory | standard (M=100, SD=15) | 5.43 / 13.31 | Yes | - | 0.35 | 0.000 | 0.21 | 0.078 | - | -2.559 | 0.343 |
|  | WISC-IV*: Digit Span | scaled (M=10, SD=3) | 1.31 / 2.69 | Yes | - | 0.38 | 0.003 | 0.25 | 0.192 | - | 0.672 | 0.620 |
|  | BRIEF-2 Parent Report: Working Memory | T-score (M=50, SD=10) | 5.26 / 9.33 | Yes | - | 0.30 | 0.050 | 0.20 | 0.476 | CHD (0.039) | 6.327 | 0.039 |
| Cognitive Flexibility | NIHTB-CB: Dimensional Card Change Sort | standard (M=100, SD=15) | 7.02 / 17.96 | No | - | 0.27 | 0.005 | 0.17 | 0.278 | Left Superior Parietal Gyrus (0.025) | -4.900 | 0.116 |
|  | D-KEFS TMT: Condition 4 - Number-Letter Switching | scaled (M=10, SD=3) | 1.19 / 3.45 | No | Left Choroid Plexus (2) | 0.42 | 0.005 | 0.20 | 0.690 | CHD (0.028) | -1.767 | 0.028 |
|  | D-KEFS VFT: Condition 3 - Category Switching | scaled (M=10, SD=3) | 1.11 / 2.99 | Yes | Left Inferior Lateral Ventricle (1), Left Lateral Ventricle (3), | 0.34 | 0.046 | 0.22 | 0.542 | Cortex - Right Hemisphere: Orbital part of the inferior frontal gyrus (0.001) | 0.000 | 1.000 |
|  |  |  |  |  | Right Inferior Lateral Ventricle (8), Right Lateral Ventricle (11) |  |  |  |  |  |  |  |
|  | D-KEFS VFT: Condition 3 - Correct Responses | scaled (M = 10, SD = 3) | 1.37 / 2.76 | Yes | Right Inferior Lateral Ventricle (10) | 0.49 | 0.000 | 0.28 | 0.202 | Cortex - Right Hemisphere: Planum polare of the superior temporal gyrus (0.022) | -0.465 | 0.538 |
|  | BRIEF-2 Parent Report: Shift | T-score (M=50, SD=10) | 5.74 / 11.25 | No | - | 0.31 | 0.034 | 0.18 | 0.639 | - | 4.392 | 0.075 |
| Inhibition | NIHTB-CB: Flanker Attention & Inhibitory Control | standard (M=100, SD=15) | 5.99 / 13.84 | Yes | - | 0.26 | 0.010 | 0.19 | 0.152 | - | -3.848 | 0.143 |
|  | D-KEFS CWIT: Condition 3 - Inhibition | scaled (M=10, SD=3) | 1.03 / 3.21 | No | Whole Brain CSF Volume (20) | 0.45 | 0.009 | 0.17 | 0.934 | - | -0.591 | 0.510 |
|  | BRIEF-2 Parent Report: Inhibit | T-score (M=50, SD=10) | 2.52 / 10.66 | No | - | 0.37 | 0.004 | 0.17 | 0.720 | - | 3.225 | 0.092 |
| Planning & Problem Solving | BRIEF-2 Parent Report: Planning & Organization | T-score (M=50, SD=10) | 4.38 / 12.36 | No | - | 0.44 | 0.000 | 0.34 | 0.015 | CHD (0.011) | 6.835 | 0.011 |

Notes. \* = 2 subjects assessed with WISC-V. Abbreviations: CWIT=Color Word Interference Test. M=Normed Mean. NIHTB-CB=NIH Toolbox Cognitive Battery. NIHTB-SB=NIH Toolbox Sensation Battery. SD=Normed Standard Deviation. TMT=Trail Making Test. VFT=Verbal Fluency Test.

#### 3.1.1 Cognitive Flexibility

For D-KEFS Trail Making Test Number-Letter Switching, we achieved a train RMSE of 1.19 and a test RMSE of 3.45. Left Choroid Plexus volume was ranked 2 out of the top 20 features in this model (Figure 2). Additionally, CSF-related structures in top 20 were proximate projection and association white matter tracts that straddle the left lateral ventricle, including the left cingulum (paralimbic cortical association-radial diffusivity-rank 1), the left cortical spinal tract (primary motor projection-fractional anisotropy), the left superior longitudinal fasciculus-(cortical association-fractional anisotropy) and the left arcuate fasciculus (cortical association fibers-fractional anisotropy), selected cortical grey matter structures, and more distal contralateral subcortical structures (right thalamus, right cerebellum and right hippocampus) Regression model with top 20 features had R2 =0.42 and Left Choroid Plexus had an estimated parameter of −0.00003247, and p=0.967. In LASSO regression R2=0.204, and Left Choroid Plexus had an estimated parameter of 0.0003, and p=0.706.

For D-KEFS Verbal Fluency Test Category Switching, we achieve train RMSE of 1.11 and test RMSE of 2.99, which was within performance threshold. Left Inferior Lateral Ventricle (ranked 1), Left Lateral Ventricle (ranked 3), Right Inferior Lateral Ventricle (ranked 8), and Right Lateral Ventricle (ranked 11) were among the top 20 features in this model (Figure 3). The proximate projection and association white matter tracts that straddle the left lateral ventricle, including the bilateral cingulum (paralimbic cortical association-radial diffusivity), the bilateral cortical spinal tract (primary motor-axial diffusivity), the bilateral superior longitudinal fasiculus (cortical association-radial diffusivity) and bilateral frontal occipital longitudinal fasciculus-(cortical association-fractional anisotropy) and the bilateral arcuate fasciculus (cortical association fibers-fractional anisotropy), selected cortical grey matter structures, and subcortical structures (basal ganglia – caudate and pallidum). Additionally, CSF-related structures in top 20 were middle posterior corpus callosum (rank 13) and left caudate (rank 14) volumes. Besides the CSF and CSF related structures, the rest of the top 20 features were composed of 8 cortical association fibers, 2 cortical grey matter structures, and 1 subcortical structure. Regression model with top 20 features had R2 =0.342, and the four CSF volume features among the top 20 had the following results: Right Lateral Ventricle had an estimated parameter of −0.0002, and p=0.24; Right Inferior Lateral Ventricle had an estimated parameter of −0.0011, and p=0.53; Left Lateral Ventricle had an estimated parameter of 0.0003, and p=0.123; and Left Inferior Lateral Ventricle had an estimated parameter of −0.002, and p=0.354. In LASSO regression R2=0.22, and the four CSF volume features among the top 20 had the following results: Right Lateral Ventricle had an estimated parameter of −6.435e-06, and p=0.976; Right Inferior Lateral Ventricle had an estimated parameter of −0.0026, and p=0.179; Left Lateral Ventricle had an estimated parameter of 8.834e-05, and p=0.708; and Left Inferior Lateral Ventricle had an estimated parameter of −0.001, and p=0.665.

**Figure 3.**
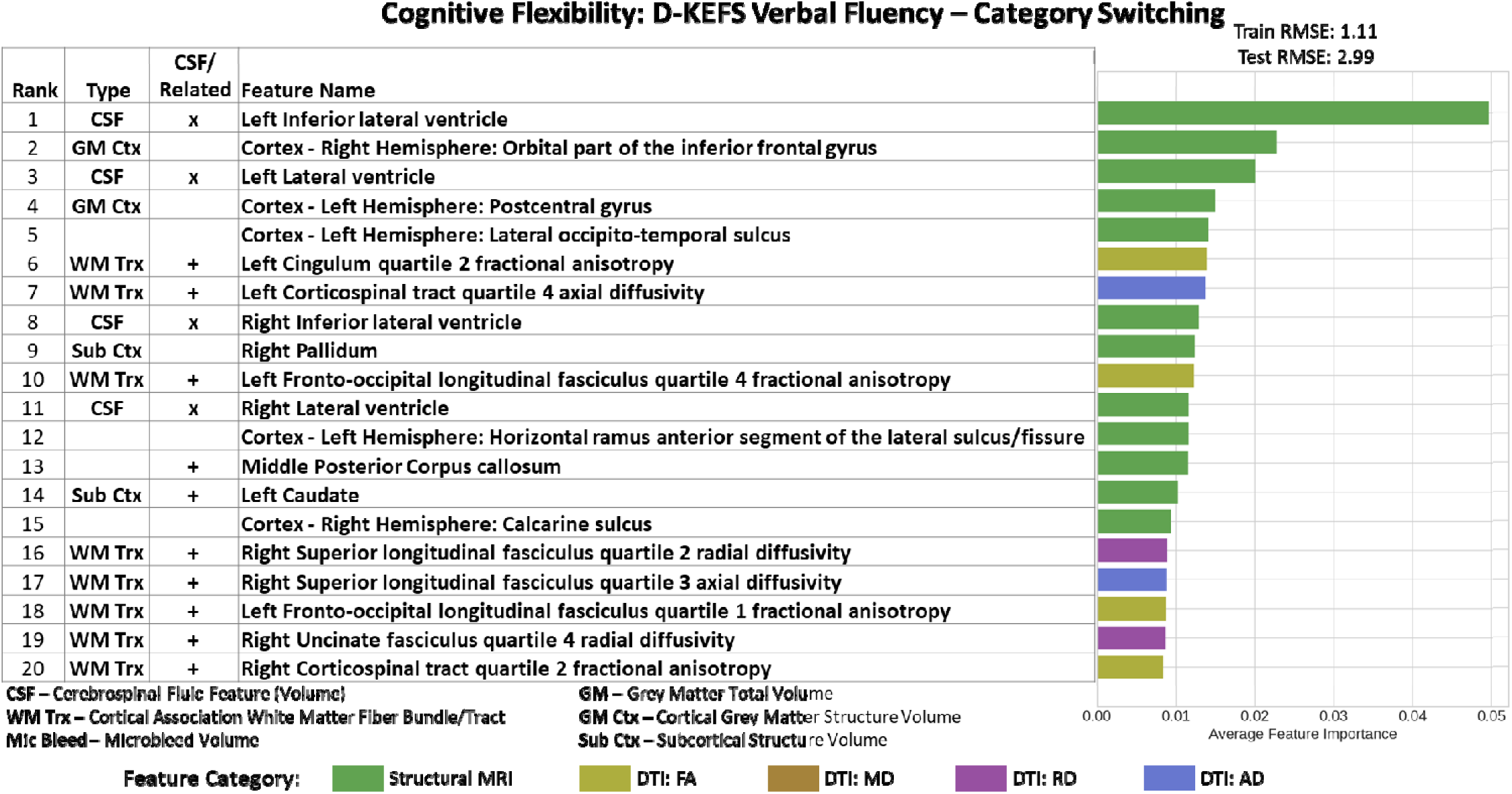
Imaging Only Top 20 Feature Importance results for Cognitive Flexibility Domain assessed using D-KEFS Verbal Fluency Category Switching Test. Left Inferior Lateral Ventricle (rank 1), Left Lateral Ventricle (rank 3), Right Inferior Lateral Ventricle (rank 8), and Right Lateral Ventricle (rank 11) were the important CSF feature in this model. The proximate projection and association white matter tracts that straddle the left lateral ventricle, including the bilateral cingulum (paralimbic cortical association-radial diffusivity), the bilateral cortical spinal tract (primary motor-axial diffusivity), the bilateral superior longitudinal fasiculus (cortical association-radial diffusivity) and bilateral frontal occipital longitudinal fasciculus-(cortical association-fractional anisotropy) and the bilateral arcuate fasciculus (cortical association fibers-fractional anisotropy), selected cortical grey matter structures, and subcortical structures (basal ganglia – caudate and pallidum).

For D-KEFS Verbal Fluency Correct Responses, we achieved train RMSE of 1.37 and test RMSE of 2.76, which was within performance threshold. Right Inferior Lateral Ventricle volumes was ranked 10 out of the top 20 features in this model (Figure 4). The proximate association white matter tracts that straddle the right lateral ventricle, including the right cingulum (paralimbic cortical association-radial diffusivity), the right fronto-occipital fasciculus-(cortical association-axial diffusivity) and the right uncinate fasciculus (cortical association axial diffusivity). Additional important imaging features include microbleed volume. There were no structures related to the right inferior lateral ventricle among the other top 20 features. Besides the CSF feature, the rest of the top 20 features were composed of 10 cortical association fibers, 6 cortical grey matter structures, 1 subcortical structure, and non-white matter microbleed volume. Regression model with top 20 features had R2 =0.489 and Right Inferior Lateral Ventricle had an estimated parameter of −0.0018, and p=0.037. In LASSO regression R2=0.281, and Right Inferior Lateral Ventricle had an estimated parameter of −0.0014, and p=0.168.

**Figure 4.**
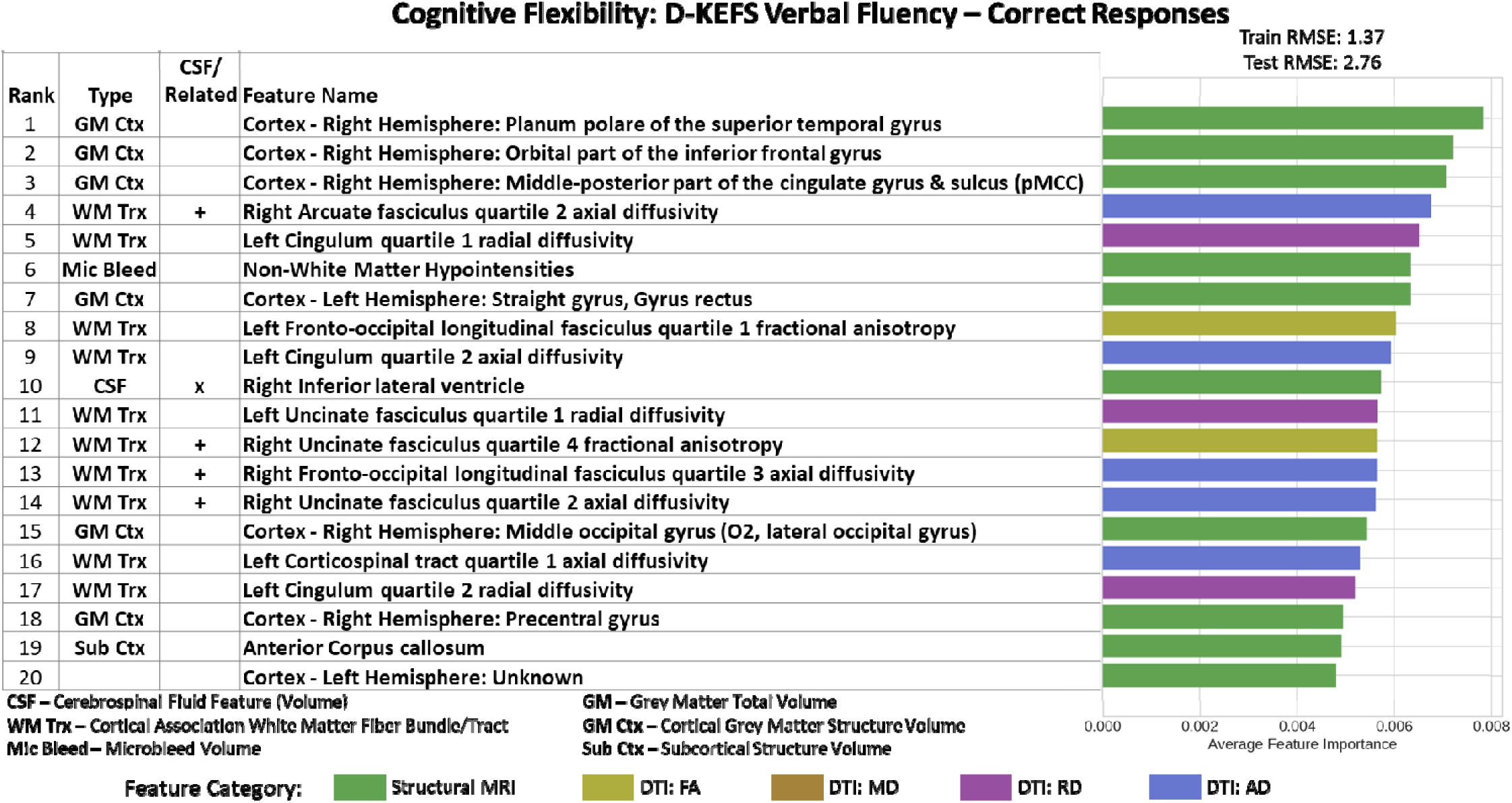
Imaging Only Top 20 Feature Importance results for Cognitive Flexibility Domain assessed using D-KEFS Verbal Fluency Category Correct Responses Test. Right Inferior Lateral Ventricle (rank 10) was the important CSF feature in this model. The proximate association white matter tracts that straddle the right lateral ventricle, including the right cingulum (paralimbic cortical association-radial diffusivity), the right fronto-occipital fasciculus-(cortical association-axial diffusivity) and the right uncinate fasciculus (cortical associationaxial diffusivity) Additional important imaging features include microbleed volume.

#### 3.1.2 Working Memory

For BRIEF 2 Working Memory, we achieved train RMSE of 5.26 and test RMSE of 9.33, which was within performance threshold. There were no CSF findings among the top 20 features in this model. In LASSO regression R2=0.20, CHD status was the only significant feature (p=0.039) with having CHD predicting poorer outcomes.

#### 3.1.3 Inhibition

For D-KEFS CWIT Inhibition, we achieved train RMSE of 1.03 and test RMSE of 3.21. Whole Brain CSF (PVE) volume was ranked 20 out of the top 20 features in this model (Figure 5). Since the entire CSF volume was an important feature in this model, potentially, many of the structures throughout the inner and outer surface of the brain tissue could potentially be considered CSF related features. Additional important imaging features that straddle the ventricular system include multiple proximate cortical projections (bilateral cortical association fibers) and cortical association fibers (cingulum, superior longitudinal fasciculus, arcuate fasciculus, uncinate fasciculus), subcortical structures (right caudate), and microbleed volumes. The rest of the top 20 features were composed of 12 cortical association fibers, 2 subcortical structures, 2 cortical grey matter structure, and both white matter and non-white matter microbleed volumes. In LASSO regression, there were no significant associations between top 20 features and outcome.

**Figure 5.**
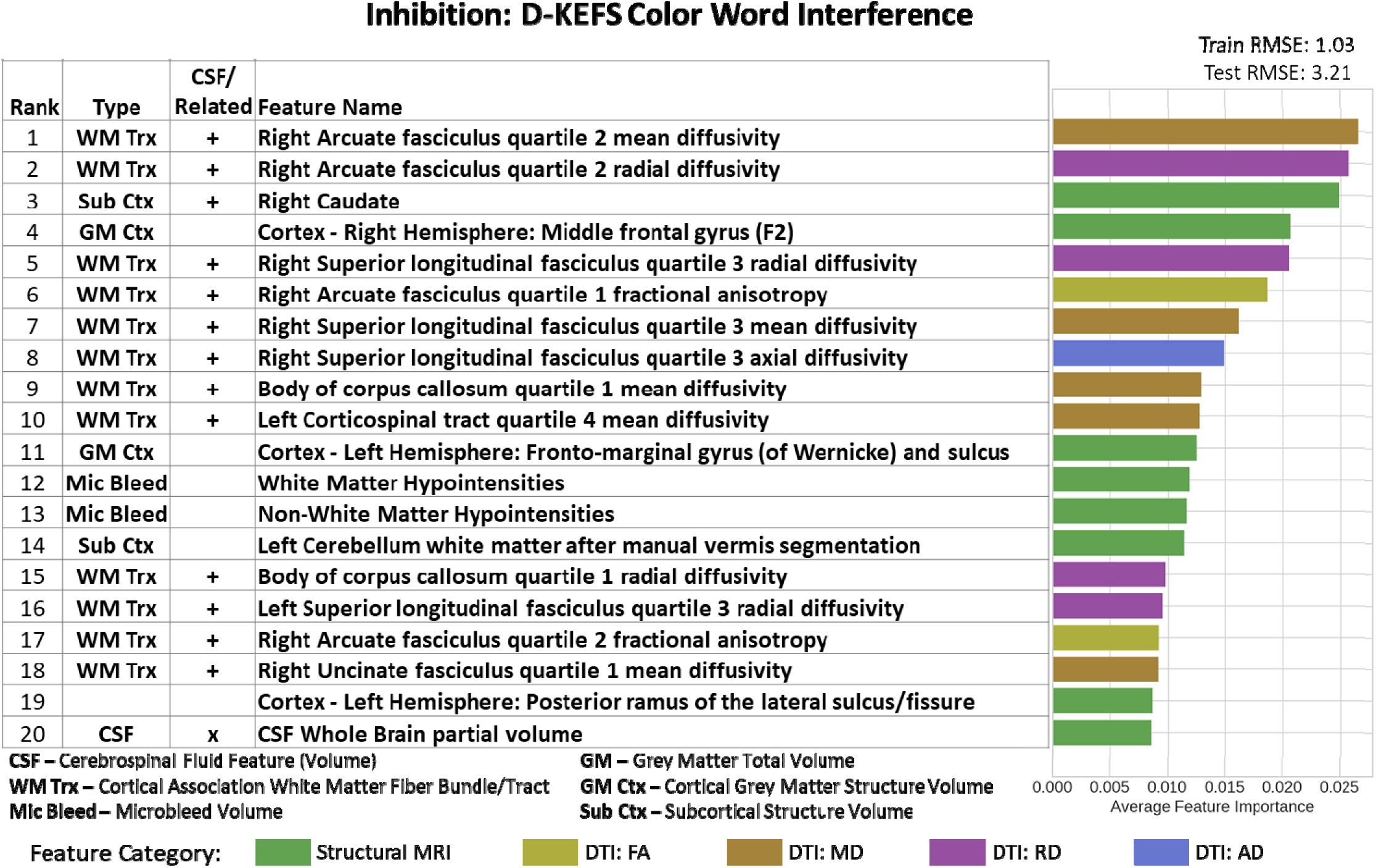
Imaging Only Top 20 Feature Importance results for Inhibition Domain assessed using D-KEFS Color Word Interference Test. Whole brain CSF volume (rank 20) was the important CSF feature in this model. Additional important imaging features that straddle the ventricular system include multiple proximate cortical projections (bilateral cortical association fibers)_ and cortical association fibers (cingulum, superior longitudinal fasciculus, arcuate fasciculus, uncinate fasciculus), subcortical structures (right caudate), and microbleed volumes.

#### 3.1.4 Planning and Problem Solving

For BRIEF-2 Planning and Organization, we achieved train RMSE of 4.38 and test RMSE of 12.36. There were no CSF findings among the top 20 features in this model. In LASSO regression R2=0.34, CHD status was the only significant feature (p=0.011) with having CHD predicting poorer outcomes.

### 3.2 Random Forest Clinical Features Model of Executive Function

The list of clinical features is in Supplemental Table 2: Clinical (Socio-demographic, Medical, Surgical) Features (n=104). The results from the Clinical Features – Social Determinants of Health, Medical and Surgical Risk Factors – only models are presented in Table 3 (top half).

**Table 3.**
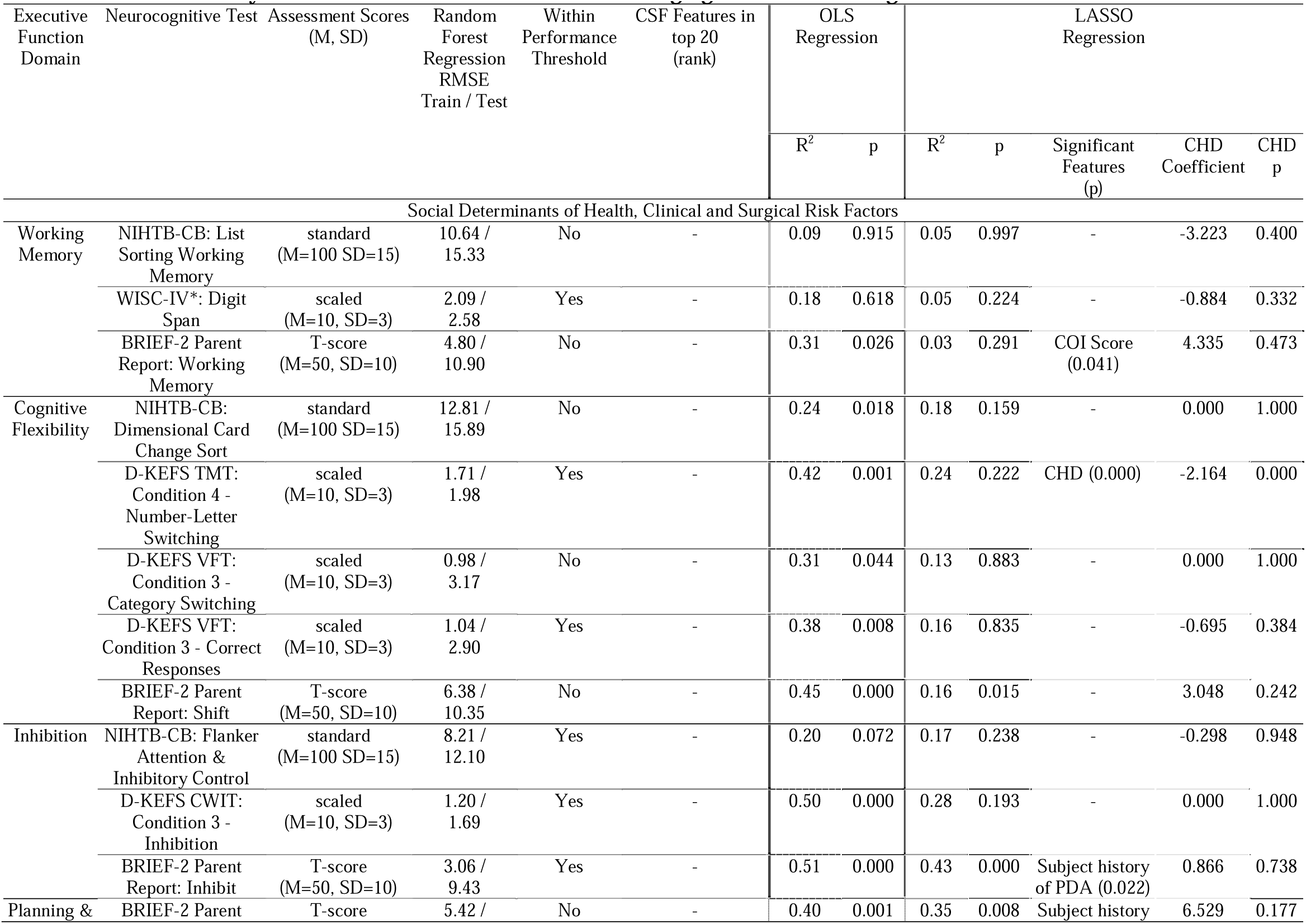
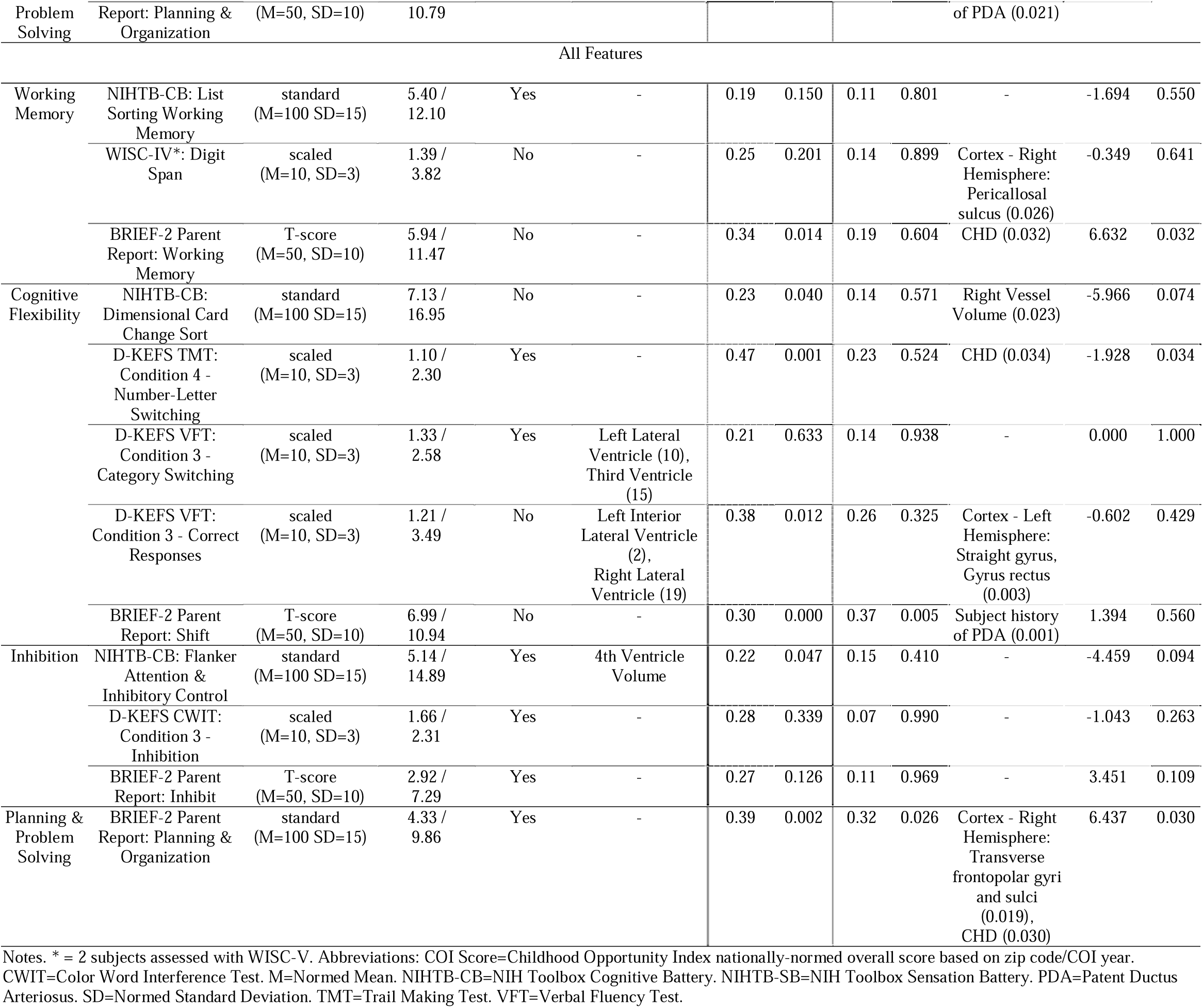
RF Clinical Only & RF Combined Clinical and Neuroimaging Model Based Regressions.

**Table 4.**
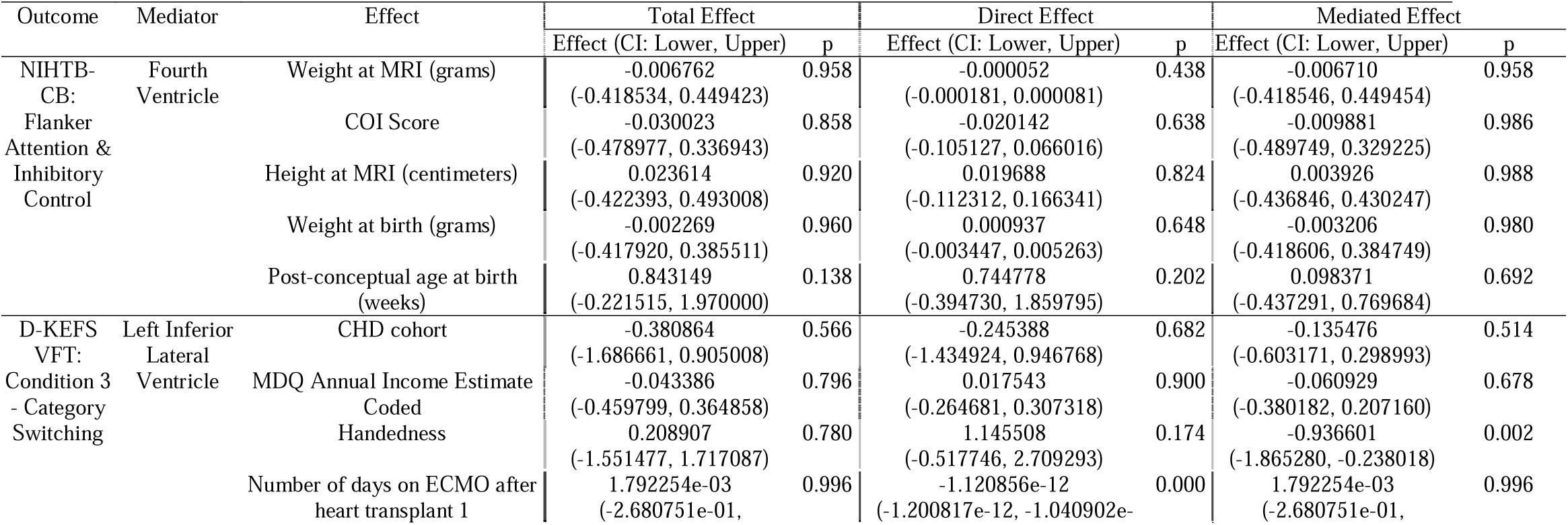
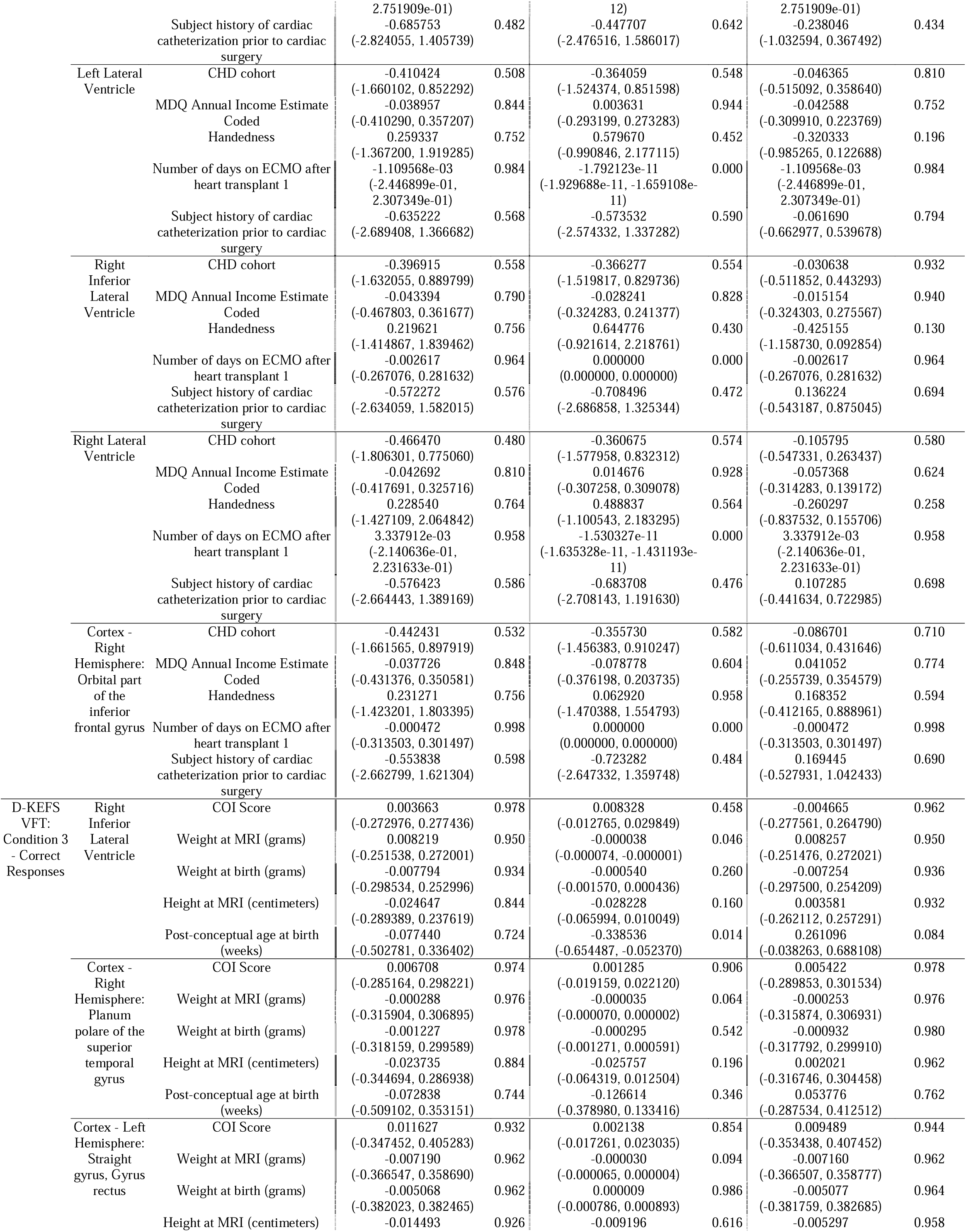
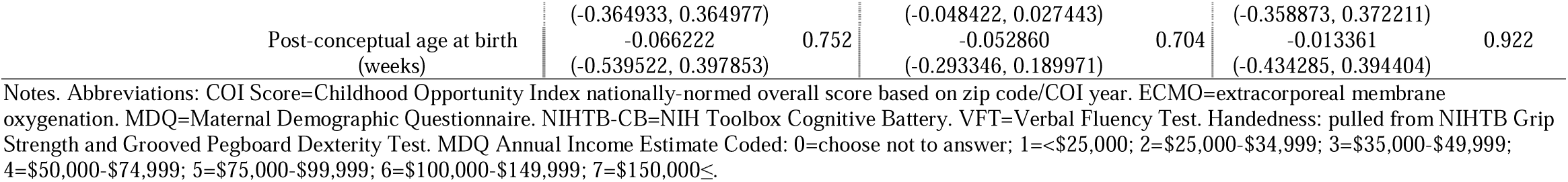
CSF Volume Related Mediated Effects.

#### 3.2.1 Cognitive Flexibility

For D-KEFS Trails Number Letter Switching, we achieved train RMSE of 1.71, test RMSE of 1.98 and achieved within performance threshold. In LASSO regression R2=0.24, and CHD status was the only significant features (p<0.001) with having CHD predicting worse outcomes.

#### 3.2.2 Working Memory

For BRIEF-2 Working Memory, we achieved train RMSE of 4.80, and test RMSE of 10.90. In LASSO regression R2=0.03, and COI was the only significant features (p=0.041), with high COI predicting better outcomes.

#### 3.2.3 Inhibition

For BRIEF-2 Inhibit, we achieved train RMSE of 3.06, and test RMSE of 9.43 and achieved within performance threshold. In LASSO regression R2=0.43, and History of PDA was the only significant features (p=0.022), with having history of PDA predicting better outcomes.

#### 3.2.4 Planning and Problem Solving

For BRIEF-2 Planning and Organization, we achieved train RMSE of 5.42, and test RMSE of 10.79. In LASSO regression R2=0.35, and History of PDA was the only significant features (p=0.021), with having history of PDA predicting better outcomes.

### 3.3 Random Forest Combined Imaging and Clinical Features Model of Executive Function

The list of all neuroimaging and clinical features in this combined model is in Supplemental Table 3: Combined Neuroimaging and Clinical Features (n=599). The results from the combined neuroimaging and clinical features (Social Determinants of Health, Medical and Surgical Risk Factors) models are presented in Table 3.

#### 3.3.1 Cognitive Flexibility

For NIHTB DCCS, we achieved train RMSE of 7.13, test RMSE of 16.95. There were no CSF findings among the top 20 features in this model. In LASSO regression (R2=0.14), right vessel volume was the only significant features (p=0.023) with larger right vessel volume predicting worse outcomes. For D-KEFS Trails Number Letter Switching, we achieved train RMSE of 1.10, test RMSE of 2.30 and achieved within performance threshold. There were no CSF findings among the top 20 features in this model. In LASSO regression R2=0.23, and CHD status was the only significant features (p<0.034) with having CHD predicting worse outcomes. For D-KEFS Verbal Fluency Switching, we achieved train RMSE of 1.33, test RMSE of 2.58 and achieved within performance threshold. Left Lateral Ventricle (rank 10), Third Ventricle (rank 15) were among the top 20 features. In LASSO regression there were no significant features associated with outcomes.

For D-KEFS Verbal Fluency Responses, we achieved train RMSE of 1.21, test RMSE of 3.49. Left Interior Lateral Ventricle (rank 2), Right Lateral Ventricle (rank 19) were among the top 20 features. In LASSO regression R2=0.26, and volume of left hemisphere gyrus rectus was the only significant features (p<0.003) with smaller volume predicting worse outcomes. For BRIEF-2 Shift, we achieved train RMSE of 6.99, and test RMSE of 10.94. There were no CSF findings among the top 20 features in this model. In LASSO regression R2=0.37, and History of PDA was the only significant features (p=0.001), with having history of PDA predicting better outcomes.

#### 3.3.2 Working Memory

For WISC-IV Digit Span, we achieved train RMSE of 1.39, test RMSE of 3.82. There were no CSF findings among the top 20 features in this model. In LASSO regression R2=0.14, Right Hemisphere Pericaollosal Sulcus volume was the only significant features (p=0.026), with having higher volume predicting worse outcomes. For BRIEF-2 Working Memory, we achieved train RMSE of 5.94, and test RMSE of 11.47. There were no CSF findings among the top 20 features in this model. In LASSO regression R2=0.19, CHD status was the only significant feature (p=0.032), with being CHD predicting poorer outcomes.

#### 3.3.3 Inhibition

There were no significant findings among the inhibition domain outcomes.

#### 3.3.4 Planning and Problem Solving

For BRIEF-2 Planning and Organization, we achieved train RMSE of 4.33, and test RMSE of 9.86 and achieved within performance threshold. There were no CSF findings among the top 20 features in this model. In LASSO regression R2=0.35, Right Hemisphere Transverse frontopolar gyri and sulci (p=0.019) and CHD status(p=0.030) were the significant features.

### 3.4 Post-Hoc Mediation and Moderation Analysis

#### 3.4.1 Post-Hoc Mediation

Post-Hoc Mediation analysis results are presented in Table 4. D-KEFS Verbal Fluency Category Switching outcome was significantly associated with the number of days of ECMO at first heart transplant in all the models. However, for this relationship, none of the imaging features selected as mediators demonstrated any significant mediation effect. Additionally, Left Inferior Lateral Ventricle Volume is mediating (p<0.01) the association between D-KEFS Verbal Fluency Category Switching outcome and NIHTB Handedness (Grooved Pegboard) main effect, although the total effect between the outcome and main effect was not statistically significant (p=0.78). D-KEFS Verbal Fluency Correct Responses outcome was significantly associated with Gestational Age (p=0.01) in the model with Right Inferior Lateral Ventricle Volume was the mediator. However, Right Inferior Lateral Ventricle Volume did not demonstrate any significant mediation effect (p=0.08).

#### 3.4.2 Post-Hoc Moderation

The Post-Hoc Moderation analysis did not demonstrated any significant total effect or moderation effect findings.

### 3.5. Feature Ontology Mapping

Table 5 shows the absolute and relative importance measures for the top 10 and top 20 features, for each random forest model of outcome using all features (imaging, clinical, surgical, demographic, and social determinants of health). The absolute feature importance measures the total contribution of each functional domain in the top 10 or 20 features identified by RFR, regardless of the proportion of features present in that functional domain. The relative feature importance shows the weighted importance of each functional domain relative to the proportion of features within that domain in the entire feature set. Therefore, features with high relative importance are likely to be more relatively impactful in predictive performance for each outcome.

**Table 5.** Feature Importance based of Feature Ontology Mapping.

| Executive Function Domain | Outcome | Within Performance Threshold | Absolute Importance: Top 10 Features |  | Relative Importance: Top 10 Features |  | Absolute Importance: Top 20 Features |  | Relative Importance: Top 20 Features |  |
| --- | --- | --- | --- | --- | --- | --- | --- | --- | --- | --- |
| Working Memory | NIHTB Cognitive Battery: List Sorting Working Memory | Yes | Paralimbic | 0.50 | Cerebellar | 0.18 | Paralimbic | 0.35 | BS | 1.00 |
|  |  |  | Cerebellar | 0.20 | Language | 0.05 | Cerebellar | 0.20 | Cerebellar | 0.36 |
|  |  |  | Visual | 0.10 | Paralimbic | 0.05 | EF | 0.10 | Language | 0.11 |
|  | WISC-IV: Digit Span | No | Language | 0.30 | Relay | 1.00 | Visual | 0.30 | Relay | 1.00 |
|  |  |  | Visual | 0.20 | Language | 0.16 | Paralimbic | 0.20 | Language | 0.21 |
|  |  |  | Paralimbic | 0.10 | GM | 0.10 | Language | 0.20 | GM | 0.20 |
|  | BRIEF-2 Parent Report: Working Memory | No | Paralimbic | 0.50 | WM | 0.25 | Paralimbic | 0.35 | DGM | 0.30 |
|  |  |  | DGM | 0.20 | DGM | 0.20 | DGM | 0.15 | WM | 0.25 |
|  |  |  | Language | 0.10 | Surgical | 0.14 | Visual | 0.15 | GM | 0.20 |
| Cognitive Flexibility | NIHTB Cognitive Battery: Dimensional Card Change Sort | No | EF | 0.30 | EF | 0.14 | EF | 0.30 | EF | 0.29 |
|  |  |  | Paralimbic | 0.20 | GM | 0.10 | Paralimbic | 0.20 | GM | 0.20 |
|  |  |  | Visual | 0.20 | CSF | 0.08 | Visual | 0.20 | CSF | 0.15 |
|  | D-KEFS Trail Making Test: Condition 4 - Number-Letter Switching | Yes | Clinical | 0.30 | Surgical | 0.14 | Visual | 0.20 | Language | 0.16 |
|  |  |  | Paralimbic | 0.20 | Language | 0.05 | Clinical | 0.20 | EF | 0.14 |
|  |  |  | Visual | 0.20 | EF | 0.05 | Paralimbic | 0.15 | Surgical | 0.14 |
|  | D-KEFS Verbal Fluency: Condition 3 - Correct Responses | No | Cerebellar | 0.40 | Cerebellar | 0.36 | Paralimbic | 0.25 | DGM | 0.40 |
|  |  |  | Paralimbic | 0.20 | DGM | 0.20 | DGM | 0.20 | Cerebellar | 0.36 |
|  |  |  | DGM | 0.20 | CSF | 0.08 | Cerebellar | 0.20 | CSF | 0.23 |
|  | D-KEFS Verbal Fluency: Condition 3 - Category Switching | Yes | Paralimbic | 0.40 | CSF | 0.15 | Paralimbic | 0.30 | Memory | 0.50 |
|  |  |  | Visual | 0.30 | Visual | 0.07 | Visual | 0.30 | CSF | 0.23 |
|  |  |  | CSF | 0.20 | EF | 0.05 | CSF | 0.15 | Visual | 0.14 |
|  | BRIEF-2 Parent Report: Shift | No | Paralimbic | 0.70 | WM | 0.25 | Paralimbic | 0.50 | WM | 0.50 |
|  |  |  | DGM | 0.10 | DGM | 0.10 | EF | 0.15 | EF | 0.14 |
|  |  |  | Clinical | 0.10 | Paralimbic | 0.07 | Clinical | 0.10 | Paralimbic | 0.10 |
| Inhibition | NIHTB Cognitive Battery: Flanker Attention & Inhibitory Control | Yes | Visual | 0.60 | Visual | 0.14 | Visual | 0.40 | Cerebellar | 0.27 |
|  |  |  | Paralimbic | 0.10 | GM | 0.10 | Paralimbic | 0.20 | Visual | 0.18 |
|  |  |  | Cerebellar | 0.10 | Cerebellar | 0.09 | Cerebellar | 0.15 | GM | 0.10 |
|  | D-KEFS Color Word Interference: Condition 3 - Inhibition | Yes | Paralimbic | 0.20 | GM | 0.10 | Paralimbic | 0.25 | CSF | 0.15 |
|  |  |  | EF | 0.20 | DGM | 0.10 | Clinical | 0.20 | EF | 0.14 |
|  |  |  | Clinical | 0.20 | EF | 0.10 | EF | 0.15 | Language | 0.11 |
|  | BRIEF-2 Parent Report: Inhibit | Yes | Paralimbic | 0.60 | Visual | 0.07 | Paralimbic | 0.50 | EF | 0.14 |
|  |  |  | Visual | 0.30 | Paralimbic | 0.06 | Visual | 0.25 | Visual | 0.11 |
|  |  |  | EF | 0.10 | EF | 0.05 | EF | 0.15 | Paralimbic | 0.10 |
| Planning and Problem Solving | BRIEF-2 Parent Report: Planning & Organization | Yes | Paralimbic | 0.40 | Cerebellar | 0.09 | Paralimbic | 0.30 | Cerebellar | 0.18 |
|  |  |  | Visual | 0.30 | Visual | 0.07 | Visual | 0.25 | Visual | 0.11 |
|  |  |  | EF | 0.10 | EF | 0.05 | EF | 0.10 | DGM | 0.10 |

Notes. Abbreviations: \* = 2 subjects assessed with WISC-V. Abbreviations: BS=brainstem. CSF=cerebrospinal fluid. CWIT=Color Word Interference Test. DGM=deep gray matter. EF=executive function. GM=gray matter. NIHTB-CB=NIH Toolbox Cognitive Battery. NIHTB-SB=NIH Toolbox Sensation Battery. TMT=Trail Making Test. VFT=Verbal Fluency Test. WM=white matter

Across all EF domains, we see a common pattern of paralimbic structures surfacing in both absolute and relative importance, and secondarily, cerebellar features being over-represented in relative importance. Working memory, particularly as measures by the NIH Toolbox List Sorting Working Memory, also showed a strong association with cerebellar and brainstem features. Cognitive flexibility outcomes showed the strongest association with executive function-brain-based domains, as well as CSF features with a strong over-representation in relative importance. Inhibition tasks had a disproportionate representation of visual systems-associated features compared to the other EF domain outcome measures.

## DISCUSSION

Studies of structure-function relationships using structural MRI in children, adolescents, and young adults with CHD have reported a variety of associations between brain integrity and outcomes.^14^ Overall smaller total and regional brain volume, as well as indicators of white matter microstructural alterations, were frequently associated with lower domain-specific cognitive outcomes, including executive functioning and memory, along with increases in ventricular sizes. ^14^ The relationship between increased CSF ventricular compartments, structural and microstructural dysmaturation, and executive function in CHD patients is unknown. Here, we leverage a novel machine-learning data-driven technique to delineate interrelationships between CSF ventricular volume, structural and microstructural alterations, clinical risk factors, and sub-domains of executive dysfunction in adolescent CHD patients. We found that CSF structural properties (including increased lateral ventricular volume and reduced choroid plexus volumes) in conjunction with proximate cortical projection and paralimbic-related association white matter tracts that straddle the lateral ventricles and distal paralimbic-related subcortical structures (basal ganglia, hippocampus, cerebellum) are predictive of two-specific subdomains of executive dysfunction in CHD patients including cognitive flexibility and inhibition. These findings in conjunction with combined RF models that incorporated clinical risk factors, highlighted important clinical risk factors, including the presence of microbleeds, altered vessel volume, and delayed PDA closure, suggesting that CSF-interstitial fluid clearance, vascular pulsatility, and glymphatic microfluid dynamics may be pathways that are impaired in congenital heart disease, providing mechanistic information about the relationship between CSF and executive dysfunctions.

The CSF serves as the medium through which water, molecules, and proteins are exchanged between brain parenchyma and vascular system, and thus, plays a critical role in regulating brain homeostasis, waste clearance, blood supply, and maintaining intracranial pressure. Prolonged interference with this clearance and homeostatic function may lead to neuroinflammation and other neurodevelopmental deficits^29^. Increased CSF volumes are seen in neonates, infants, and children with CHD^2,3,5^. Importantly, increased CSF volume has been shown to be associated with poor neurodevelopmental performance in children with CHD up to three years of age^5,6^. Likewise, disruption in CSF flow has been linked to many neurodevelopmental^30^ disorders such as autism spectrum disorder, and neurodegenerative diseases such as Alzheimer’s Disease ^31^. Our study provides support that increased CSF volumes are present in pediatric and adolescent CHD patients with specific sub-domains of executive dysfunction, including cognitive flexibility and inhibition.

A major finding of our RF models showed that CSF volumetric disturbances were associated with primate projection and association white matter tracts that straddle the lateral ventricle margins.^32^ While the classic thinking is that increase in ventricular size is secondary to central white matter damage, the findings of microbleeds (small vessel disease),^33^ altered intracranial vessel volumes and delayed PDA closure^34^ suggest that altered interstitial fluid (ISF) dynamics may be present in the CNS of CHD patients.^35^ The mechanisms driving these glymphatic fluxes have been recently described.^36^ Glymphatic system function requires adequate CSF production by the choroid plexus to provide a pressure gradient for fluid to move from the ventricles to the subarachnoid space (SAS) and subsequently into the perivascular spaces (PVS).^37–39^ The PVS is lined, almost in its entirety, by astrocytic endfeet lined with aquaporin-4 (AQP4) water channels abutting the abluminal vessel wall. Previous literature has established that AQP4 is necessary for glymphatic fluid fluxes and CSF-ISF exchange is modulated by both sleep-wake state changes and body posture. The glymphatic system is involved in several processes that become deranged in pathological states. Prior work on glymphatic failure in non-CHD small vessel disease has delineated three mechanistic pathways that could be relevant to understanding neurocognitive deficits in CHD: **(1) Perivascular spaces**, and their structural and functional integrity, have shown to be critical to homeostatic glymphatic function, specifically the role of perivascular AQP4 polarization. We propose that the CNS in CHD may exhibit vascular remodeling, abnormal perivascular spaces, and the perivascular CSF flow disturbances. **(2) Cerebrovascular pulsatility** has been demonstrated to be a driving force for CSF flow within the PVS. Extensive experimental evidence and mathematical modeling have shown tracer movements compatible with bulk flow within these spaces, although tracer entry is size-dependent and PVS blockage (i.e. protein aggregates) has shown to affect these flows. A recent theoretical model contradicts the existence of bulk flow in the PVS, but provides no supporting experimental evidence. Features of small vessel disease like changes in vascular wall compliance and reactivity are likely to alter these flows significantly, but this has not been evaluated in CHD. **(3) CNS clearance** of toxic solutes such as Aβ, has been shown to be drastically reduced in several age-related and neurodegenerative disease models, but has not also been studied in CHD, despite known dementia risk in CHD patients.. Glymphatic function has been assessed in several highly relevant processes to small vessel disease including aging and diseases such as microinfarction, stroke, Alzheimer’s disease, migraine, and diabetes. Extrapolating from the current literature, these finding suggest that CHD patients may exhibit the effects of altered PVS structure and function, arterial pulsatility, and clearance on glymphatic function.

Another major finding of our study is that CSF volumetric disturbances in adolescent CHD are associated with microstructural paralimbic (cingulum white matter tract) and subcortical paralimbic abnormalities (basal ganglia, hippocampus and cerebellum) in predicting cognitive flexibility and inhibiting executive dysfunction subdomains. Children with CHD are at a higher risk of developing brain dysmaturation, a generalized term encompassing abnormal and delayed development of brain macro- and microstructure^3,5,9,40,41^. Brain dysmaturation can take the form of qualitatively assessed brain dysplasia as detailed in our previous work^3,40^, and includes hypoplasia (reduced size) or dysplasia (malformation) of brain structures. It can also manifest quantifiably as decreased volume in regional cortical and subcortical structures^5,9,41^, decreased cortical thickness or reduced gyrification^9,42,43^, and reduced white matter microstructure or delayed myelination of white matter tracts^15,44–46^. These brain dysmaturations are associated with poor inhibitory control and executive function impairments in children and adolescents with CHD^15,47–49^. Therefore, while examining the relationship between CSF and neurodevelopment, it is essential to consider the contribution of brain dysmaturation to neurodevelopmental outcomes. Additionally, our previous studies in neonates and infants with CHD revealed the concomitant presence of brain dysmaturation and increased extra-axial CSF^3^, suggesting potential interrelationships between brain structural abnormalities and disturbed CSF properties. Normal CSF flow is believed to not only serve as nutrient distribution and waste removal^50^, but also plays an important role in neurogenesis^51^. Disruption to CSF circulation may lead to disturbance of normal neurogenesis. Previously, we have investigated whether neonates with CHD have brain dysmaturation (brain volume abnormalities, brain structure malformations, and decreased cortical thickness) of a developmental etiology independent of surgical or hypoxic brain injury^3^. Our study found that CHD was associated with olfactory bulb dysmorphometry (hypoplasia or absence), hippocampal abnormalities (dysplasia or hypoplasia), corpus callosum dysplasia, and brain stem dysplasia – which are features of brain dysmaturation^3^. Conjointly, we found that these study participants with CHD also exhibited increased extra-axial CSF volume (qualitatively assessed)^3^. In a follow-up, albeit limited study, we showed that brain dysmaturation was associated with poor neurodevelopmental outcomes^52^, which has been supported by other studies as well^10,53^

When predicting FOM, there was a strong association between model accuracy and paralimbic associated features. Prior work has shown fronto-limbic interactions, in particular amygdala-prefrontal interactions, decrease in connectivity as cognitive load increases. It is possible that the feature importance identified here could be reflective of this effect, where a dip in connectivity could be reflective of increased cognitive load and poor importance. This effect was originally found in working memory but likely extends to other cognitive domains. Additionally, we see a strong association between cerebellar and brainstem features and predictive performance in working memory tasks. This is in accordance with our prior work in adolescents with CHD showing compensatory upregulation of cerebellar pathways, and downregulation of thalamo-cortical pathways. [during which working memory tasks].^17^

Our finding of CSF volumetric disturbances, paralimbic abnormalities and right-handedness may also be related to genetic-related ciliary dysfunction.^54–56^ Despite the multitude of genes and the complexity of the interplay in genetic variation that lead to inconsistent phenotypic expression of CHD, investigations into de novo and rare inherited variants have revealed a higher burden of mutation in genes expressed in the heart or involved in heart development – and significantly in ciliary and chromatin modifying genes^57,58^. A recent large-scale mouse mutagenesis study that screened for mutations causing CHD demonstrated an important role for cilia in CHD pathogenesis^57^. This phenotype-based screen recovered over 250 mutant mouse lines with a wide spectrum of CHD, including mouse models for HLHS. These mouse models were invaluable in exposing the ways in which defects in motile ciliary genes, through disruption in cell migration, might be linked to heart defects and CHD. Motile cilia are microscopic hairlike structures found on surfaces of certain types of cells such as ependymal cells lining the epithelia surface of cerebral ventricles. Motile cilia beat in coordinated rhythmic movements to direct the flow of CSF, at least microscopically, over the ependymal surface. This movement of CSF flow is believed to contribute to nutrient distribution and waste removal^50^, as well as play an important role in neurogenesis^51^. In our prior investigation on brain dysmaturation in neonates with CHD, we found that abnormal ciliary motion was significantly associated with increased extra-axial CSF volume, as well as with olfactory bulb hypoplasia or absence, hippocampal dysplasia or hypoplasia, corpus callosum dysplasia, and brain stem dysplasia – all features of brain dysmaturation^3^. This finding of link between brain dysmaturation and abnormal ciliary motion in humans with CHD follows and supports similar well characterized findings in CHD mouse model. The mouse model also demonstrated motile cilia’s important role in CHD pathogenesis^57^. Along with our study, findings of brain abnormalities and intellectual disability in ciliopathies, such as Bardet-Biedl syndrome^59^, suggest that dysfunctional motile cilia may play a role in sequelae of brain dysmaturation in patients with CHD.

Neurodevelopmental disabilities in CHD population have not been fully explained by the multitude of research examining genetics, epigenetic, and clinical risk factors. The association between ciliary motion dysfunction, CSF disruption, and neurodevelopmental disabilities in CHD have yet to be explored together.

### Limitations

Using Gini coefficient for deriving feature importance has some limitations. When two features are highly correlated, the Gini importance might be distributed unevenly among them, attributing higher importance to one and neglecting the other, even though they might equally contribute to the predictive power. We mitigate this by using FOM to identify broader feature categories contributing to classification accuracy. Additionally, although Gini importance provides a global measure of feature importance, it doesn’t necessarily offer comprehensive insight into how a feature influences predictions across all possible values and contexts within the model. Finally, random forests are inherently stochastic, making the absolute importance ranking of any given feature prone to some random variation. This is mitigated by looking at a fixed set of top features (i.e. top 20 features) and validation feature importance with more rigorous statistical methods and feature selection. While feature ontology mapping is a powerful tool for enhancing model interpretability and domain relevance, it is essential to be mindful of some limitations and challenges. Ensuring that the ontology accurately represents all necessary domain knowledge remains a challenge for future work. It may not be feasible to capture all relevant domain knowledge within the ontology, leading to potential knowledge gaps. Additionally, some features have multiple functional associations, or unspecified functions. We attempted to map each feature to a canonically and functionally useful primary association. Future work should attempt to assign weighted importance in multi-association ontologies.

### Future Directions

Future work can examine other features of CSF including flow. The flow of CSF can be characterized as bulk flow and pulsatile flow, with the former describing the overall circulation and absorption of the CSF while the latter describes the dynamic flow of CSF through the ventricles and cisterns driven by cardiopulmonary circulatory processes^60^. The synchronicity of CSF flow to cardiac cycle means that any abnormal cardiac function or injury in the cardiovascular system – common comorbidities of CHD^61^ – that could disrupt cerebral blood flow could also adversely affect CSF flow and ultimately CSF circulation. Thus, examining the pulsatile CSF flow would be ideal to further elucidate the possible connections between the abnormal hemodynamics endemic in CHD population and CSF flow characteristics. Future work can quantify CSF flow in vivo, using phase contrast MRI (PCMRI), in children and adolescents with CHD. Other future work could focus on mapping the glymphatic or microfluid system which can be assessed with diffusion MR, perfusion MR and MR elastography.^36^

### Conclusion

Here, we leverage a novel machine-learning data-driven technique to delineate interrelationships between CSF ventricular volume, structural and microstructural alterations, clinical risk factors, and sub-domains of executive dysfunction in adolescent CHD patients. We found that CSF structural properties (including increased lateral ventricular volume and reduced choroid plexus volumes) in conjunction with proximate cortical projection and paralimbic-related association white matter tracts that straddle the lateral ventricles and distal paralimbic-related subcortical structures (basal ganglia, hippocampus, cerebellum) are predictive of two-specific subdomains of executive dysfunction in CHD patients including cognitive flexibility and inhibition. These findings in conjunction with combined RF models that incorporated clinical risk factors, highlighted important clinical risk factors, including the presence of microbleeds, altered vessel volume, and delayed PDA closure, suggesting that CSF-interstitial fluid clearance, vascular pulsatility, and glymphatic microfluid dynamics may be pathways that are impaired in congenital heart disease, providing mechanistic information about the relationship between CSF and executive dysfunctions.

## Data Availability

All data produced in the present study are available upon reasonable request to the authors.

## Notes

This work was supported by the Department of Defense (W81XWH-16-1-0613), the National Heart, Lung and Blood Institute (R01 HL152740-1 and R01 HL128818-05), the National Heart, Lung and Blood Institute with National Institute of Aging (R01 HL128818-05 S1), the National Institute of Neurological Disorders and Stroke (K23 063371), the National Library of Medicine (5T15LM007059-27) and Additional Ventures.

### Competing Interest Statement

The authors have declared no competing interest.

### Funding Statement

This study was funded by the Department of Defense (W81XWH-16-1-0613), the National Heart, Lung and Blood Institute (R01 HL152740-1 and R01 HL128818-05), the National Heart, Lung and Blood Institute with National Institute of Aging (R01 HL128818-05 S1), the National Institute of Neurological Disorders and Stroke (K23 063371), the National Library of Medicine (5T15LM007059-27) and Additional Ventures.

### Author Declarations

Ethics committee/IRB of The University of Pittsburgh (University of Pittsburgh Institutional Review Board) gave ethical approval for this work. Consent was obtained from the parent/guardian of all participants.

