## Supplement for "Cerebral Spinal Fluid Volumetrics and Paralimbic Predictors of Executive Dysfunction in Congenital Heart Disease: A Machine Learning Approach Informing Mechanistic Insights"

**Supplement Material**

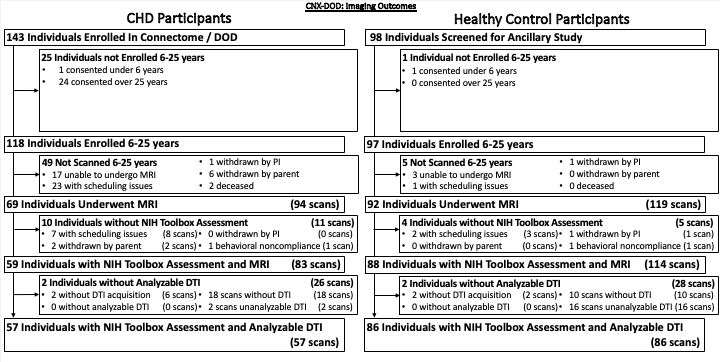

**Supplemental Figure 1: Recruitment Flow Diagram**

**Supplemental Table 1: Neuroimaging Features (n= 495)**

| **Feature (s)** | **N-Features** |
| --- | --- |
| Brain Stem Volume | 1 |
| Inferior Vermis Volume | 1 |
| Left/Right Cerebellum Cortex | 2 |
| Left/Right Cerebellum White Matter | 2 |
| Manual Vermis Segmentation | 1 |
| Middle Vermis Volume | 1 |
| Superior Vermis Volume | 1 |
| 3rd Ventricle Volume | 1 |
| 4th Ventricle Volume | 1 |
| 5th Ventricle Volume | 1 |
| Left/Right Choroid Plexus Volume | 2 |
| Left/Right Inferior Lateral Ventricle Volume | 2 |
| Left/Right Lateral Ventricle Volume | 2 |
| Left/Right Ventral DC Volume | 2 |
| Left/Right Vessel Volume | 2 |
| Optic Chiasm Volume | 1 |
| Total CSF Volume (discrete volume model) | 1 |
| Total CSF Volume (Intracranial) | 1 |
| Total CSF Volume (partial volume model) | 1 |
| Left/Right Accumbens Area Volume | 2 |
| Left/Right Caudate Volume | 2 |
| Left/Right Pallidum Volume | 2 |
| Left/Right Putamen Volume | 2 |
| Left/Right Thalamus Proper Volume | 2 |
| Left/Right Frontal Middle Sulcus Volume | 2 |
| Left/Right Frontomarginal Gyrus and Sulcus vulume | 2 |
| Left/Right Gyrus Rectus Volume | 2 |
| Left/Right Inferior Frontoal Opercular Gyrus Volume | 2 |
| Left/Right Superior Frontal Gyrus Volume | 2 |
| Left/Right Superior Frontal Sulcus Volume | 2 |
| Left/Right Uncinate Fasciculus (Q1-4) FA, MD, RD, AD | 32 |
| Left/Right Precuneus Gyrus Volume | 2 |
| Freesurfer: Left/Right Unclassified Cortex Volume | 2 |
| Left/Right Anterior Horizontal Lateral Fissure Volume | 2 |
| Left/Right Anterior Vertical Lateral Fissure Volume | 2 |
| Left/Right Cerebral Cortex Volume | 2 |
| Left/Right Posterior Lateral Fissure Volume | 2 |
| Total Grey Matter Volume (discrete volume model) | 1 |
| Total Grey Matter Volume (partial volume model) | 1 |
| Left/Right Temporal-Superior Lateral Gyrus | 2 |
| Left/Right Arcuate Fasciculus (Q1-4) FA, MD, RD, AD | 32 |
| Left/Right Fronto-polar Transverse Gyrus and Sulcus Vulume | 2 |
| Left/Right Inferior Temporal Gyrus Volume | 2 |
| Left/Right Inferior Temporal Sulcus Volume | 2 |
| Left/Right Inferior-Frontal Triangular Gyrus Volume | 2 |
| Left/Right Inferior-Parietal Supramarginal Gyrus Volume | 2 |
| Left/Right Middle Frontal Gyrus Volume | 2 |
| Left/Right Middle Temporal Gyrus Volume | 2 |
| Left/Right Subcentral Gyrus and Sulcus Volume | 2 |
| Left/Right Superior Longitudinal Fasciculus (Q1-4) FA, MD, RD, AD | 32 |
| Left/Right Superior Temporal Sulcus Volume | 2 |
| Left/Right Temporal Polar Gyrus Volume | 2 |
| Left/Right Temporal Pole Volume | 2 |
| Left/Right Transverse Temporal Sulcus Volume | 2 |
| Left/Right Anterior Circular Insular Sulcus Volume | 2 |
| Left/Right Central Insular Gyrus and Sulcus Volume | 2 |
| Left/Right Central Sulcus Volume | 2 |
| Left/Right Corticospinal Tract (Q1-4) FA, MD, RD, AD | 32 |
| Left/Right Inferior Circular Insular Sulcus Volume | 2 |
| Left/Right Inferior Perietal Precentral Sulcus Volume | 2 |
| Left/Right Intermediate Pirmnary Jensen Sulcus Volume | 2 |
| Left/Right Paracentral Gyrus and Sulcus Volume | 2 |
| Left/Right Postcentral Gyrus Volume | 2 |
| Left/Right Postcentral Sulcus Volume | 2 |
| Left/Right Posteror-Transverse Intraparietal Sulcus Volume | 2 |
| Left/Right precentral Gyrus Volume | 2 |
| Left/Right Short Insular Gyrus Volume | 2 |
| Left/Right Subcallosal Gyrus Volume | 2 |
| Left/Right Superior Circular Insular Sulcus Volume | 2 |
| Left/Right Superior Parietal Precentral Sulcus Volume | 2 |
| Left/Right Subparietral Sulcus Volume | 2 |
| Body, Genu, Splenium of Corpus Callosum (Q1-4) FA, MD, RD, AD | 48 |
| Corpus Callosum: Anterior, Mid-Anterior, Central, Posterior Volume | 4 |
| Fornix (Q1-4) FA, MD, RD, AD | 16 |
| Left/Right Anterior, Middle, Mid-Posterior, Posterior-Dorsal, Posterior-Ventral Cingulate Gyrus and Sulcus Volume | 10 |
| Left/Right Cingular Marginalis Sulcus Volume | 2 |
| Left/Right Cingulum (Q1-4) FA, MD, RD, AD | 32 |
| Left/Right Hippocampus Volume | 2 |
| Left/Right H-Shaped Orbital Sulcus Volume | 2 |
| Left/Right Inferior Frontal Sulcus Volume | 2 |
| Left/Right Inferior Fronto-Orbital Gyrus Volume | 2 |
| Left/Right Inferior-Parietal Angular Gyrus Volume | 2 |
| Left/Right Lateral Orbital Sulcus Volume | 2 |
| Left/Right Medial Parahippocampal Gyrus Volume | 2 |
| Left/Right Medial-Orbital Olfactory Sulcus Volume | 2 |
| Left/Right Orbital Gyrus Volume | 2 |
| Left/Right Pericallosal Sulcus Volume | 2 |
| Left/Right Suborbital Sulcus Volume | 2 |
| Left/Right Amygdala Volume | 2 |
| Left/Right Manual Hippocampus Segmentation | 2 |
| Left/Right Anterior Collateral Transverse Sulcus Volume | 2 |
| Left/Right Anterior Occipital Sulcus Volume | 2 |
| Left/Right Calcarine Sulcus Volume | 2 |
| Left/Right Cuneus Gyrus Volume | 2 |
| Left/Right Fronto-Occipital Fasciculus (Q1-4) FA, MD, RD, AD | 32 |
| Left/Right Inferior Longitudinal Fasciculus (Q1-4) FA, MD, RD, AD | 32 |
| Left/Right Inferior Occipital Gyrus and Sulcus Volume | 2 |
| Left/Right Lateral Occipital Sulcus Volume | 2 |
| Left/Right Medial Temporal and Lingual Suclus Volume | 2 |
| left/Right Medial-Lingual Occipital Temporal Gyrus Volume | 2 |
| Left/Right Middle Occipital Gyrus Volume | 2 |
| Left/Right Occipital Middle and Lunatus Sulcus Volume | 2 |
| Left/Right Occipital Parietal Sulcus Volume | 2 |
| Left/Right Occipital Pole Volume | 2 |
| Left/Right Occipital-Temporal Fusiform Gyrus Volume | 2 |
| Left/Right Posterior Collateral Transverse Sulcus Volume | 2 |
| Left/Right Superior Occipital and Transversal Sulcus Volume | 2 |
| Left/Right Superior Occipital Gyrus Volume | 2 |
| Left/Right Superior Parietal Gyrus Volume | 2 |
| Left/Right Temporal Pole Volume | 2 |
| Left/Right Temporal-Superior Transverse Gyrus Volume | 2 |
| Whole Brain Volume | 1 |
| Left/Right Cerebral White Matter Volume | 2 |
| Non-WM Hypointensities Volume | 1 |
| Total White Matter Volume (discrete volume model) | 1 |
| Total White Matter Volume (partial volume model) | 1 |
| WM Hypointensities Volume | 1 |
| TOTAL FEATURES | 495 |

**Supplemental Table 2: Clinical (Socio-demographic, Medical, Surgical) Features (n=104)**

| **Feature (s)** | **N-Features** |
| --- | --- |
| Age at Scan (YOS-YOB) | 1 |
| Sex | 1 |
| Maternal Education | 1 |
| Age at Assessment | 1 |
| ECMO | 1 |
| From Norwood to Fontan, Total number of catheterizations | 1 |
| Hospital LOS (Stage I) | 1 |
| Hospital LOS (Stage II) | 1 |
| Hospital LOS (Stage III) | 1 |
| Hospital LOS (Summation of Stages II and III) | 1 |
| Norwood, ECMO used during hospitalization | 1 |
| Norwood, ECMO used in OR | 1 |
| Pre-Norwood, Number of cath interventions | 1 |
| Total number of cardiac catheterizations | 1 |
| Aortic Arch Repair | 1 |
| DORV Repair | 1 |
| Pulmonary Valve Repair | 1 |
| Ross-Konno | 1 |
| Tricupid Valve Repair | 1 |
| Truncus A. Repair | 1 |
| Total anomalous pulmonary venous return repair | 1 |
| Tetralogy of Fallot Repair | 1 |
| Right Ventricle to Pulmonary Artery Conduit Repair | 1 |
| Right Ventricular Outflow Tract Repair | 1 |
| Sinus Venous Atrial Septal Defect Repair | 1 |
| AVSD-VSD-ASD-PFO-PDA Closure | 1 |
| Other Intervention | 1 |
| Number of Heart Transplants | 1 |
| Total Length of Stay | 1 |
| Surgical Length of Stay | 1 |
| ECMO Total Days | 1 |
| Age at First Complication | 1 |
| Age at Last Seizure | 1 |
| Age at Last Stroke | 1 |
| Anatomy, Genetic syndrome | 1 |
| CHD | 1 |
| Clinical Event Ever Through 12 Years, Stroke | 1 |
| From Norwood to Fontan, Total number of complications | 1 |
| Heart Transplant Date (First Transplant) | 1 |
| LBW (<2.5kg) | 1 |
| Number of unplanned reinterventions | 1 |
| Prematurity | 1 |
| Pre-Norwood, Number of complications | 1 |
| Primary indication for intervention | 1 |
| Seizure (dichotomized as any/none) | 1 |
| Seizure since last contact | 1 |
| Stroke (dichotomized as any/none) | 1 |
| Stroke since last contact | 1 |
| Through 12 months, Number of Serious Adverse Events | 1 |
| Total Number of Serior Adverse Events | 1 |
| Gestational Age | 1 |
| Birth Weight (g) | 1 |
| Birthweight (less than 2.5g) | 1 |
| Occipito-Frontal Head Circumference at Birth (cm) | 1 |
| Length at Birth (cm) | 1 |
| Seizure | 1 |
| Age at Onset of First Seizure | 1 |
| Stroke | 1 |
| Number of Strokes | 1 |
| Infarct | 1 |
| Hemorrhage | 1 |
| Encephalomalacia | 1 |
| Ventriculomegaly/Hydrocephalus | 1 |
| Gliosis | 1 |
| Necrosis | 1 |
| Periventricular Leukomalacia (PVL) | 1 |
| Volume Loss on MRI | 1 |
| Structural Abnormalities | 1 |
| Number Ventricles | 1 |
| Cyanosis | 1 |
| Left ventricular outflow tract obstruction (LVOTO) | 1 |
| Single Ventricle | 1 |
| Hypoplastic Left Heart Syndrome | 1 |
| Hypoplastic Right Ventricle/ Hypoplastic Left Ventricle | 1 |
| Ebsteins | 1 |
| Shones | 1 |
| Double Outlet Right Ventricle | 1 |
| Double Inlet Left Ventricle | 1 |
| Transposition of the Great Aorta | 1 |
| Tetralogy of Fallot | 1 |
| Tricuspid Atresia | 1 |
| Pulmonary Valve Atresia | 1 |
| Truncus Arteriosis | 1 |
| Partial/Total anomalous pulmonary venous return ( | 1 |
| Aortic Abnormality | 1 |
| Coarctation of Aorta | 1 |
| Hypoplastic Aortic Arch | 1 |
| Interrupted Aortic Arch | 1 |
| Bicuspid Aortic Arch | 1 |
| Aortic Atresia | 1 |
| Aortic Stenosis | 1 |
| Dextrocardia | 1 |
| Atrioventricular Septal Defect | 1 |
| Pulmonary Stenosis | 1 |
| Mitral Atresia | 1 |
| Mitral Stenosis | 1 |
| Ventricular Septal Defect | 1 |
| Atrial Septal Defect | 1 |
| Patent Ductus Arteriosus | 1 |
| Patent Foramen Ovale | 1 |
| Cardiac Catheterization | 1 |
| Number of Cardiac Catheterizations | 1 |
| Total Lifetime Surgeries | 1 |
| Age at First Surgery | 1 |
| TOTAL FEATURES | 104 |

**Supplemental Table 3: Combined Neuroimaging and Clinical Features (n=599)**

| **Feature (s)** | **N-Features** |
| --- | --- |
| Brain Stem Volume | 1 |
| Inferior Vermis Volume | 1 |
| Left/Right Cerebellum Cortex | 2 |
| Left/Right Cerebellum White Matter | 2 |
| Manual Vermis Segmentation | 1 |
| Middle Vermis Volume | 1 |
| Superior Vermis Volume | 1 |
| 3rd Ventricle Volume | 1 |
| 4th Ventricle Volume | 1 |
| 5th Ventricle Volume | 1 |
| Left/Right Choroid Plexus Volume | 2 |
| Left/Right Inferior Lateral Ventricle Volume | 2 |
| Left/Right Lateral Ventricle Volume | 2 |
| Left/Right Ventral DC Volume | 2 |
| Left/Right Vessel Volume | 2 |
| Optic Chiasm Volume | 1 |
| Total CSF Volume (discrete volume model) | 1 |
| Total CSF Volume (Intracranial) | 1 |
| Total CSF Volume (partial volume model) | 1 |
| Left/Right Accumbens Area Volume | 2 |
| Left/Right Caudate Volume | 2 |
| Left/Right Pallidum Volume | 2 |
| Left/Right Putamen Volume | 2 |
| Left/Right Thalamus Proper Volume | 2 |
| Left/Right Frontal Middle Sulcus Volume | 2 |
| Left/Right Frontomarginal Gyrus and Sulcus vulume | 2 |
| Left/Right Gyrus Rectus Volume | 2 |
| Left/Right Inferior Frontoal Opercular Gyrus Volume | 2 |
| Left/Right Superior Frontal Gyrus Volume | 2 |
| Left/Right Superior Frontal Sulcus Volume | 2 |
| Left/Right Uncinate Fasciculus (Q1-4) FA, MD, RD, AD | 32 |
| Left/Right Precuneus Gyrus Volume | 2 |
| Freesurfer: Left/Right Unclassified Cortex Volume | 2 |
| Left/Right Anterior Horizontal Lateral Fissure Volume | 2 |
| Left/Right Anterior Vertical Lateral Fissure Volume | 2 |
| Left/Right Cerebral Cortex Volume | 2 |
| Left/Right Posterior Lateral Fissure Volume | 2 |
| Total Grey Matter Volume (discrete volume model) | 1 |
| Total Grey Matter Volume (partial volume model) | 1 |
| Left/Right Temporal-Superior Lateral Gyrus | 2 |
| Left/Right Arcuate Fasciculus (Q1-4) FA, MD, RD, AD | 32 |
| Left/Right Fronto-polar Transverse Gyrus and Sulcus Vulume | 2 |
| Left/Right Inferior Temporal Gyrus Volume | 2 |
| Left/Right Inferior Temporal Sulcus Volume | 2 |
| Left/Right Inferior-Frontal Triangular Gyrus Volume | 2 |
| Left/Right Inferior-Parietal Supramarginal Gyrus Volume | 2 |
| Left/Right Middle Frontal Gyrus Volume | 2 |
| Left/Right Middle Temporal Gyrus Volume | 2 |
| Left/Right Subcentral Gyrus and Sulcus Volume | 2 |
| Left/Right Superior Longitudinal Fasciculus (Q1-4) FA, MD, RD, AD | 32 |
| Left/Right Superior Temporal Sulcus Volume | 2 |
| Left/Right Temporal Polar Gyrus Volume | 2 |
| Left/Right Temporal Pole Volume | 2 |
| Left/Right Transverse Temporal Sulcus Volume | 2 |
| Left/Right Anterior Circular Insular Sulcus Volume | 2 |
| Left/Right Central Insular Gyrus and Sulcus Volume | 2 |
| Left/Right Central Sulcus Volume | 2 |
| Left/Right Corticospinal Tract (Q1-4) FA, MD, RD, AD | 32 |
| Left/Right Inferior Circular Insular Sulcus Volume | 2 |
| Left/Right Inferior Perietal Precentral Sulcus Volume | 2 |
| Left/Right Intermediate Pirmnary Jensen Sulcus Volume | 2 |
| Left/Right Paracentral Gyrus and Sulcus Volume | 2 |
| Left/Right Postcentral Gyrus Volume | 2 |
| Left/Right Postcentral Sulcus Volume | 2 |
| Left/Right Posteror-Transverse Intraparietal Sulcus Volume | 2 |
| Left/Right precentral Gyrus Volume | 2 |
| Left/Right Short Insular Gyrus Volume | 2 |
| Left/Right Subcallosal Gyrus Volume | 2 |
| Left/Right Superior Circular Insular Sulcus Volume | 2 |
| Left/Right Superior Parietal Precentral Sulcus Volume | 2 |
| Left/Right Subparietral Sulcus Volume | 2 |
| Body, Genu, Splenium of Corpus Callosum (Q1-4) FA, MD, RD, AD | 48 |
| Corpus Callosum: Anterior, Mid-Anterior, Central, Posterior Volume | 4 |
| Fornix (Q1-4) FA, MD, RD, AD | 16 |
| Left/Right Anterior, Middle, Mid-Posterior, Posterior-Dorsal, Posterior-Ventral Cingulate Gyrus and Sulcus Volume | 10 |
| Left/Right Cingular Marginalis Sulcus Volume | 2 |
| Left/Right Cingulum (Q1-4) FA, MD, RD, AD | 32 |
| Left/Right Hippocampus Volume | 2 |
| Left/Right H-Shaped Orbital Sulcus Volume | 2 |
| Left/Right Inferior Frontal Sulcus Volume | 2 |
| Left/Right Inferior Fronto-Orbital Gyrus Volume | 2 |
| Left/Right Inferior-Parietal Angular Gyrus Volume | 2 |
| Left/Right Lateral Orbital Sulcus Volume | 2 |
| Left/Right Medial Parahippocampal Gyrus Volume | 2 |
| Left/Right Medial-Orbital Olfactory Sulcus Volume | 2 |
| Left/Right Orbital Gyrus Volume | 2 |
| Left/Right Pericallosal Sulcus Volume | 2 |
| Left/Right Suborbital Sulcus Volume | 2 |
| Left/Right Amygdala Volume | 2 |
| Left/Right Manual Hippocampus Segmentation | 2 |
| Left/Right Anterior Collateral Transverse Sulcus Volume | 2 |
| Left/Right Anterior Occipital Sulcus Volume | 2 |
| Left/Right Calcarine Sulcus Volume | 2 |
| Left/Right Cuneus Gyrus Volume | 2 |
| Left/Right Fronto-Occipital Fasciculus (Q1-4) FA, MD, RD, AD | 32 |
| Left/Right Inferior Longitudinal Fasciculus (Q1-4) FA, MD, RD, AD | 32 |
| Left/Right Inferior Occipital Gyrus and Sulcus Volume | 2 |
| Left/Right Lateral Occipital Sulcus Volume | 2 |
| Left/Right Medial Temporal and Lingual Suclus Volume | 2 |
| left/Right Medial-Lingual Occipital Temporal Gyrus Volume | 2 |
| Left/Right Middle Occipital Gyrus Volume | 2 |
| Left/Right Occipital Middle and Lunatus Sulcus Volume | 2 |
| Left/Right Occipital Parietal Sulcus Volume | 2 |
| Left/Right Occipital Pole Volume | 2 |
| Left/Right Occipital-Temporal Fusiform Gyrus Volume | 2 |
| Left/Right Posterior Collateral Transverse Sulcus Volume | 2 |
| Left/Right Superior Occipital and Transversal Sulcus Volume | 2 |
| Left/Right Superior Occipital Gyrus Volume | 2 |
| Left/Right Superior Parietal Gyrus Volume | 2 |
| Left/Right Temporal Pole Volume | 2 |
| Left/Right Temporal-Superior Transverse Gyrus Volume | 2 |
| Whole Brain Volume | 1 |
| Left/Right Cerebral White Matter Volume | 2 |
| Non-WM Hypointensities Volume | 1 |
| Total White Matter Volume (discrete volume model) | 1 |
| Total White Matter Volume (partial volume model) | 1 |
| WM Hypointensities Volume | 1 |
| Age at Scan (YOS-YOB) | 1 |
| Sex | 1 |
| Maternal Education | 1 |
| Age at Assessment | 1 |
| ECMO | 1 |
| From Norwood to Fontan, Total number of catheterizations | 1 |
| Hospital LOS (Stage I) | 1 |
| Hospital LOS (Stage II) | 1 |
| Hospital LOS (Stage III) | 1 |
| Hospital LOS (Summation of Stages II and III) | 1 |
| Norwood, ECMO used during hospitalization | 1 |
| Norwood, ECMO used in OR | 1 |
| Pre-Norwood, Number of cath interventions | 1 |
| Total number of cardiac catheterizations | 1 |
| Aortic Arch Repair | 1 |
| DORV Repair | 1 |
| Pulmonary Valve Repair | 1 |
| Ross-Konno | 1 |
| Tricupid Valve Repair | 1 |
| Truncus A. Repair | 1 |
| Total anomalous pulmonary venous return repair | 1 |
| Tetralogy of Fallot Repair | 1 |
| Right Ventricle to Pulmonary Artery Conduit Repair | 1 |
| Right Ventricular Outflow Tract Repair | 1 |
| Sinus Venous Atrial Septal Defect Repair | 1 |
| AVSD-VSD-ASD-PFO-PDA Closure | 1 |
| Other Intervention | 1 |
| Number of Heart Transplants | 1 |
| Total Length of Stay | 1 |
| Surgical Length of Stay | 1 |
| ECMO Total Days | 1 |
| Age at First Complication | 1 |
| Age at Last Seizure | 1 |
| Age at Last Stroke | 1 |
| Anatomy, Genetic syndrome | 1 |
| CHD | 1 |
| Clinical Event Ever Through 12 Years, Stroke | 1 |
| From Norwood to Fontan, Total number of complications | 1 |
| Heart Transplant Date (First Transplant) | 1 |
| LBW (<2.5kg) | 1 |
| Number of unplanned reinterventions | 1 |
| Prematurity | 1 |
| Pre-Norwood, Number of complications | 1 |
| Primary indication for intervention | 1 |
| Seizure (dichotomized as any/none) | 1 |
| Seizure since last contact | 1 |
| Stroke (dichotomized as any/none) | 1 |
| Stroke since last contact | 1 |
| Through 12 months, Number of Serious Adverse Events | 1 |
| Total Number of Serior Adverse Events | 1 |
| Gestational Age | 1 |
| Birth Weight (g) | 1 |
| Birthweight (less than 2.5g) | 1 |
| Occipito-Frontal Head Circumference at Birth (cm) | 1 |
| Length at Birth (cm) | 1 |
| Seizure | 1 |
| Age at Onset of First Seizure | 1 |
| Stroke | 1 |
| Number of Strokes | 1 |
| Infarct | 1 |
| Hemorrhage | 1 |
| Encephalomalacia | 1 |
| Ventriculomegaly/Hydrocephalus | 1 |
| Gliosis | 1 |
| Necrosis | 1 |
| Periventricular Leukomalacia (PVL) | 1 |
| Volume Loss on MRI | 1 |
| Structural Abnormalities | 1 |
| Number Ventricles | 1 |
| Cyanosis | 1 |
| Left ventricular outflow tract obstruction (LVOTO) | 1 |
| Single Ventricle | 1 |
| Hypoplastic Left Heart Syndrome | 1 |
| Hypoplastic Right Ventricle/ Hypoplastic Left Ventricle | 1 |
| Ebsteins | 1 |
| Shones | 1 |
| Double Outlet Right Ventricle | 1 |
| Double Inlet Left Ventricle | 1 |
| Transposition of the Great Aorta | 1 |
| Tetralogy of Fallot | 1 |
| Tricuspid Atresia | 1 |
| Pulmonary Valve Atresia | 1 |
| Truncus Arteriosis | 1 |
| Partial/Total anomalous pulmonary venous return ( | 1 |
| Aortic Abnormality | 1 |
| Coarctation of Aorta | 1 |
| Hypoplastic Aortic Arch | 1 |
| Interrupted Aortic Arch | 1 |
| Bicuspid Aortic Arch | 1 |
| Aortic Atresia | 1 |
| Aortic Stenosis | 1 |
| Dextrocardia | 1 |
| Atrioventricular Septal Defect | 1 |
| Pulmonary Stenosis | 1 |
| Mitral Atresia | 1 |
| Mitral Stenosis | 1 |
| Ventricular Septal Defect | 1 |
| Atrial Septal Defect | 1 |
| Patent Ductus Arteriosus | 1 |
| Patent Foramen Ovale | 1 |
| Cardiac Catheterization | 1 |
| Number of Cardiac Catheterizations | 1 |
| Total Lifetime Surgeries | 1 |
| Age at First Surgery | 1 |
| TOTAL FEATURES | 599 |

**Supplemental Table 4: Feature Ontology Mapping Classification**

| **Feature (s)** | **Domain** | **N-Features** |
| --- | --- | --- |
| Brain Stem Volume | BS | 1 |
| Inferior Vermis Volume | Cerebellar | 1 |
| Left/Right Cerebellum Cortex | Cerebellar | 2 |
| Left/Right Cerebellum White Matter | Cerebellar | 2 |
| Manual Vermis Segmentation | Cerebellar | 1 |
| Middle Vermis Volume | Cerebellar | 1 |
| Superior Vermis Volume | Cerebellar | 1 |
| 3rd Ventricle Volume | CSF | 1 |
| 4th Ventricle Volume | CSF | 1 |
| 5th Ventricle Volume | CSF | 1 |
| Left/Right Choroid Plexus Volume | CSF | 2 |
| Left/Right Inferior Lateral Ventricle Volume | CSF | 2 |
| Left/Right Lateral Ventricle Volume | CSF | 2 |
| Left/Right Ventral DC Volume | CSF | 2 |
| Left/Right Vessel Volume | CSF | 2 |
| Optic Chiasm Volume | CSF | 1 |
| Total CSF Volume (discrete volume model) | CSF | 1 |
| Total CSF Volume (Intracranial) | CSF | 1 |
| Total CSF Volume (partial volume model) | CSF | 1 |
| Left/Right Accumbens Area Volume | DGM | 2 |
| Left/Right Caudate Volume | DGM | 2 |
| Left/Right Pallidum Volume | DGM | 2 |
| Left/Right Putamen Volume | DGM | 2 |
| Left/Right Thalamus Proper Volume | DGM | 2 |
| Left/Right Frontal Middle Sulcus Volume | EF | 2 |
| Left/Right Frontomarginal Gyrus and Sulcus vulume | EF | 2 |
| Left/Right Gyrus Rectus Volume | EF | 2 |
| Left/Right Inferior Frontoal Opercular Gyrus Volume | EF | 2 |
| Left/Right Superior Frontal Gyrus Volume | EF | 2 |
| Left/Right Superior Frontal Sulcus Volume | EF | 2 |
| Left/Right Uncinate Fasciculus (Q1-4) FA, MD, RD, AD | EF | 32 |
| Left/Right Precuneus Gyrus Volume | EF | 2 |
| Freesurfer: Left/Right Unclassified Cortex Volume | GM | 2 |
| Left/Right Anterior Horizontal Lateral Fissure Volume | GM | 2 |
| Left/Right Anterior Vertical Lateral Fissure Volume | GM | 2 |
| Left/Right Cerebral Cortex Volume | GM | 2 |
| Left/Right Posterior Lateral Fissure Volume | GM | 2 |
| Total Grey Matter Volume (discrete volume model) | GM | 1 |
| Total Grey Matter Volume (partial volume model) | GM | 1 |
| Left/Right Temporal-Superior Lateral Gyrus | Language | 2 |
| Left/Right Arcuate Fasciculus (Q1-4) FA, MD, RD, AD | Language | 32 |
| Left/Right Fronto-polar Transverse Gyrus and Sulcus Vulume | Language | 2 |
| Left/Right Inferior Temporal Gyrus Volume | Language | 2 |
| Left/Right Inferior Temporal Sulcus Volume | Language | 2 |
| Left/Right Inferior-Frontal Triangular Gyrus Volume | Language | 2 |
| Left/Right Inferior-Parietal Supramarginal Gyrus Volume | Language | 2 |
| Left/Right Middle Frontal Gyrus Volume | Language | 2 |
| Left/Right Middle Temporal Gyrus Volume | Language | 2 |
| Left/Right Subcentral Gyrus and Sulcus Volume | Language | 2 |
| Left/Right Superior Longitudinal Fasciculus (Q1-4) FA, MD, RD, AD | Language | 32 |
| Left/Right Superior Temporal Sulcus Volume | Language | 2 |
| Left/Right Temporal Polar Gyrus Volume | Language | 2 |
| Left/Right Temporal Pole Volume | Language | 2 |
| Left/Right Transverse Temporal Sulcus Volume | Language | 2 |
| Left/Right Anterior Circular Insular Sulcus Volume | Motor | 2 |
| Left/Right Central Insular Gyrus and Sulcus Volume | Motor | 2 |
| Left/Right Central Sulcus Volume | Motor | 2 |
| Left/Right Corticospinal Tract (Q1-4) FA, MD, RD, AD | Motor | 32 |
| Left/Right Inferior Circular Insular Sulcus Volume | Motor | 2 |
| Left/Right Inferior Perietal Precentral Sulcus Volume | Motor | 2 |
| Left/Right Intermediate Pirmnary Jensen Sulcus Volume | Motor | 2 |
| Left/Right Paracentral Gyrus and Sulcus Volume | Motor | 2 |
| Left/Right Postcentral Gyrus Volume | Motor | 2 |
| Left/Right Postcentral Sulcus Volume | Motor | 2 |
| Left/Right Posteror-Transverse Intraparietal Sulcus Volume | Motor | 2 |
| Left/Right precentral Gyrus Volume | Motor | 2 |
| Left/Right Short Insular Gyrus Volume | Motor | 2 |
| Left/Right Subcallosal Gyrus Volume | Motor | 2 |
| Left/Right Superior Circular Insular Sulcus Volume | Motor | 2 |
| Left/Right Superior Parietal Precentral Sulcus Volume | Motor | 2 |
| Left/Right Subparietral Sulcus Volume | Paralimbic | 2 |
| Body, Genu, Splenium of Corpus Callosum (Q1-4) FA, MD, RD, AD | Paralimbic | 48 |
| Corpus Callosum: Anterior, Mid-Anterior, Central, Posterior Volume | Paralimbic | 4 |
| Fornix (Q1-4) FA, MD, RD, AD | Paralimbic | 16 |
| Left/Right Anterior, Middle, Mid-Posterior, Posterior-Dorsal, Posterior-Ventral Cingulate Gyrus and Sulcus Volume | Paralimbic | 10 |
| Left/Right Cingular Marginalis Sulcus Volume | Paralimbic | 2 |
| Left/Right Cingulum (Q1-4) FA, MD, RD, AD | Paralimbic | 32 |
| Left/Right Hippocampus Volume | Paralimbic | 2 |
| Left/Right H-Shaped Orbital Sulcus Volume | Paralimbic | 2 |
| Left/Right Inferior Frontal Sulcus Volume | Paralimbic | 2 |
| Left/Right Inferior Fronto-Orbital Gyrus Volume | Paralimbic | 2 |
| Left/Right Inferior-Parietal Angular Gyrus Volume | Paralimbic | 2 |
| Left/Right Lateral Orbital Sulcus Volume | Paralimbic | 2 |
| Left/Right Medial Parahippocampal Gyrus Volume | Paralimbic | 2 |
| Left/Right Medial-Orbital Olfactory Sulcus Volume | Paralimbic | 2 |
| Left/Right Orbital Gyrus Volume | Paralimbic | 2 |
| Left/Right Pericallosal Sulcus Volume | Paralimbic | 2 |
| Left/Right Suborbital Sulcus Volume | Paralimbic | 2 |
| Left/Right Amygdala Volume | Paralimbic | 2 |
| Left/Right Manual Hippocampus Segmentation | Paralimbic | 2 |
| Left/Right Anterior Collateral Transverse Sulcus Volume | Visual | 2 |
| Left/Right Anterior Occipital Sulcus Volume | Visual | 2 |
| Left/Right Calcarine Sulcus Volume | Visual | 2 |
| Left/Right Cuneus Gyrus Volume | Visual | 2 |
| Left/Right Fronto-Occipital Fasciculus (Q1-4) FA, MD, RD, AD | Visual | 32 |
| Left/Right Inferior Longitudinal Fasciculus (Q1-4) FA, MD, RD, AD | Visual | 32 |
| Left/Right Inferior Occipital Gyrus and Sulcus Volume | Visual | 2 |
| Left/Right Lateral Occipital Sulcus Volume | Visual | 2 |
| Left/Right Medial Temporal and Lingual Suclus Volume | Visual | 2 |
| left/Right Medial-Lingual Occipital Temporal Gyrus Volume | Visual | 2 |
| Left/Right Middle Occipital Gyrus Volume | Visual | 2 |
| Left/Right Occipital Middle and Lunatus Sulcus Volume | Visual | 2 |
| Left/Right Occipital Parietal Sulcus Volume | Visual | 2 |
| Left/Right Occipital Pole Volume | Visual | 2 |
| Left/Right Occipital-Temporal Fusiform Gyrus Volume | Visual | 2 |
| Left/Right Posterior Collateral Transverse Sulcus Volume | Visual | 2 |
| Left/Right Superior Occipital and Transversal Sulcus Volume | Visual | 2 |
| Left/Right Superior Occipital Gyrus Volume | Visual | 2 |
| Left/Right Superior Parietal Gyrus Volume | Visual | 2 |
| Left/Right Temporal Pole Volume | Visual | 2 |
| Left/Right Temporal-Superior Transverse Gyrus Volume | Visual | 2 |
| Whole Brain Volume | Whole Brain | 1 |
| Left/Right Cerebral White Matter Volume | WM | 2 |
| Non-WM Hypointensities Volume | WM | 1 |
| Total White Matter Volume (discrete volume model) | WM | 1 |
| Total White Matter Volume (partial volume model) | WM | 1 |
| WM Hypointensities Volume | WM | 1 |
| Age at Scan (YOS-YOB) | Demographic | 1 |
| Sex | Demographic | 1 |
| Maternal Education | Demographic | 1 |
| Age at Assessment | Demographic | 1 |
| ECMO | Surgical | 1 |
| From Norwood to Fontan, Total number of catheterizations | Surgical | 1 |
| Hospital LOS (Stage I) | Surgical | 1 |
| Hospital LOS (Stage II) | Surgical | 1 |
| Hospital LOS (Stage III) | Surgical | 1 |
| Hospital LOS (Summation of Stages II and III) | Surgical | 1 |
| Norwood, ECMO used during hospitalization | Surgical | 1 |
| Norwood, ECMO used in OR | Surgical | 1 |
| Pre-Norwood, Number of cath interventions | Surgical | 1 |
| Total number of cardiac catheterizations | Surgical | 1 |
| Aortic Arch Repair | Surgical | 1 |
| DORV Repair | Surgical | 1 |
| Pulmonary Valve Repair | Surgical | 1 |
| Ross-Konno | Surgical | 1 |
| Tricupid Valve Repair | Surgical | 1 |
| Truncus A. Repair | Surgical | 1 |
| Total anomalous pulmonary venous return repair | Surgical | 1 |
| Tetralogy of Fallot Repair | Surgical | 1 |
| Right Ventricle to Pulmonary Artery Conduit Repair | Surgical | 1 |
| Right Ventricular Outflow Tract Repair | Surgical | 1 |
| Sinus Venous Atrial Septal Defect Repair | Surgical | 1 |
| AVSD-VSD-ASD-PFO-PDA Closure | Surgical | 1 |
| Other Intervention | Surgical | 1 |
| Number of Heart Transplants | Surgical | 1 |
| Total Length of Stay | Surgical | 1 |
| Surgical Length of Stay | Surgical | 1 |
| ECMO Total Days | Surgical | 1 |
| Age at First Complication | Clinical | 1 |
| Age at Last Seizure | Clinical | 1 |
| Age at Last Stroke | Clinical | 1 |
| Anatomy, Genetic syndrome | Clinical | 1 |
| CHD | Clinical | 1 |
| Clinical Event Ever Through 12 Years, Stroke | Clinical | 1 |
| From Norwood to Fontan, Total number of complications | Clinical | 1 |
| Heart Transplant Date (First Transplant) | Clinical | 1 |
| LBW (<2.5kg) | Clinical | 1 |
| Number of unplanned reinterventions | Clinical | 1 |
| Prematurity | Clinical | 1 |
| Pre-Norwood, Number of complications | Clinical | 1 |
| Primary indication for intervention | Clinical | 1 |
| Seizure (dichotomized as any/none) | Clinical | 1 |
| Seizure since last contact | Clinical | 1 |
| Stroke (dichotomized as any/none) | Clinical | 1 |
| Stroke since last contact | Clinical | 1 |
| Through 12 months, Number of Serious Adverse Events | Clinical | 1 |
| Total Number of Serior Adverse Events | Clinical | 1 |
| Gestational Age | Clinical | 1 |
| Birth Weight (g) | Clinical | 1 |
| Birthweight (less than 2.5g) | Clinical | 1 |
| Occipito-Frontal Head Circumference at Birth (cm) | Clinical | 1 |
| Length at Birth (cm) | Clinical | 1 |
| Seizure | Clinical | 1 |
| Age at Onset of First Seizure | Clinical | 1 |
| Stroke | Clinical | 1 |
| Number of Strokes | Clinical | 1 |
| Infarct | Clinical | 1 |
| Hemorrhage | Clinical | 1 |
| Encephalomalacia | Clinical | 1 |
| Ventriculomegaly/Hydrocephalus | Clinical | 1 |
| Gliosis | Clinical | 1 |
| Necrosis | Clinical | 1 |
| Periventricular Leukomalacia (PVL) | Clinical | 1 |
| Volume Loss on MRI | Clinical | 1 |
| Structural Abnormalities | Clinical | 1 |
| Number Ventricles | Clinical | 1 |
| Cyanosis | Clinical | 1 |
| Left ventricular outflow tract obstruction (LVOTO) | Clinical | 1 |
| Single Ventricle | Clinical | 1 |
| Hypoplastic Left Heart Syndrome | Clinical | 1 |
| Hypoplastic Right Ventricle/ Hypoplastic Left Ventricle | Clinical | 1 |
| Ebsteins | Clinical | 1 |
| Shones | Clinical | 1 |
| Double Outlet Right Ventricle | Clinical | 1 |
| Double Inlet Left Ventricle | Clinical | 1 |
| Transposition of the Great Aorta | Clinical | 1 |
| Tetralogy of Fallot | Clinical | 1 |
| Tricuspid Atresia | Clinical | 1 |
| Pulmonary Valve Atresia | Clinical | 1 |
| Truncus Arteriosis | Clinical | 1 |
| Partial/Total anomalous pulmonary venous return ( | Clinical | 1 |
| Aortic Abnormality | Clinical | 1 |
| Coarctation of Aorta | Clinical | 1 |
| Hypoplastic Aortic Arch | Clinical | 1 |
| Interrupted Aortic Arch | Clinical | 1 |
| Bicuspid Aortic Arch | Clinical | 1 |
| Aortic Atresia | Clinical | 1 |
| Aortic Stenosis | Clinical | 1 |
| Dextrocardia | Clinical | 1 |
| Atrioventricular Septal Defect | Clinical | 1 |
| Pulmonary Stenosis | Clinical | 1 |
| Mitral Atresia | Clinical | 1 |
| Mitral Stenosis | Clinical | 1 |
| Ventricular Septal Defect | Clinical | 1 |
| Atrial Septal Defect | Clinical | 1 |
| Patent Ductus Arteriosus | Clinical | 1 |
| Patent Foramen Ovale | Clinical | 1 |
| Cardiac Catheterization | Clinical | 1 |
| Number of Cardiac Catheterizations | Clinical | 1 |
| Total Lifetime Surgeries | Clinical | 1 |
| Age at First Surgery | Clinical | 1 |
